# Unlocking the Global Antigenic Diversity and Balancing Selection of *Plasmodium falciparum*

**DOI:** 10.1101/2021.06.21.21259065

**Authors:** Myo T. Naung, Elijah Martin, Jacob Munro, Somya Mehra, Andrew J. Guy, Moses Laman, G.L. Abby Harrison, Livingstone Tavul, Manuel Hetzel, Dominic Kwiatkowski, Ivo Mueller, Melanie Bahlo, Alyssa E. Barry

## Abstract

Investigation of the diversity of malaria parasite antigens can help prioritize and validate them as vaccine candidates and identify the most common variants for inclusion in vaccine formulations. Studies on *Plasmodium falciparum* antigen diversity have focused on well-known vaccine candidates while the diversity of several others has never been studied. Here we provide an overview of the diversity and population structure of leading vaccine candidate antigens of *P. falciparum* using the MalariaGEN Pf3K (version 5.1) resource, comprising more than 2600 genomes from 15 malaria endemic countries. We developed a stringent variant calling pipeline to extract high quality antigen gene sequences from the global dataset and a new R-package named *VaxPack* to streamline population genetic analyses. In addition, a newly developed algorithm that enables spatial averaging of selection pressure on 3D protein structures was applied to the dataset. We analysed the genes encoding 23 leading and novel candidate malaria vaccine antigens including *csp*, *trap*, *eba175*, *ama1*, *rh5*, and *CelTOS*. We found that current malaria vaccine formulations are based on rare variants and thus may have limited efficacy. High levels of diversity with evidence of balancing selection was detected for most of the erythrocytic and pre-erythrocytic antigens. Measures of natural selection were then mapped to 3D protein structures to predict targets of functional antibodies. For some antigens, geographical variation in the intensity and distribution of these signals on the 3D structure suggests adaptations to different human host or mosquito vector populations. This study provides an essential framework for the diversity of *P. falciparum* antigens for inclusion in the design of the next generation of malaria vaccines.

**Author Summary:** Highly effective malaria vaccines are important for the sustainable elimination of malaria. However, the diversity of parasite antigens targeted by malaria vaccines has been largely overlooked, with most vaccine formulations based only on a single antigen variant. Failure to accommodate this diversity may result in vaccines only being effective against vaccine-like variants, resulting in limited protective efficacy. Investigation of the diversity of genes encoding parasite antigens can help prioritize and validate them as vaccine candidates as well as to identify the most common variants for inclusion in the next generation of malaria vaccines. Here we measure the diversity of 23 vaccine antigens of *Plasmodium falciparum*, using the publicly available MalariaGEN Pf3K (version 5.1) resource comprising more than 2600 genomes from 15 malaria endemic countries. We found that variants found in current vaccine formulations are rare and thus may target only a small proportion of circulating malaria parasite strains. Variation in intensity of immune selection in parasites from different geographic areas suggests adaptation to different human host or vector populations. This study provides an essential framework for the design of the next generation of malaria vaccines, in addition to providing novel insights into malaria biology.

## Introduction

*Plasmodium falciparum*, the most lethal human malaria parasite, has been co-evolving with its human host for tens of thousands of years [1]. Along this evolutionary timeline, host-parasite interactions have left landmarks of selection on both genomes [2, 3]. These landmarks can help to identify parasite surface proteins targeted by host immune responses and therefore antigens that could be used in “subunit” vaccines to protect against infection and disease. However, malaria parasites evade host immune responses through the accumulation of mutations, thus increasing the repertoire of variants circulating in the parasite population, and therefore this diversity needs to be considered in vaccine formulations [5, 6]. Otherwise, vaccines may only be partially effective resulting in a “sieve effect”, with higher efficacy against vaccine-like strains and low to no efficacy against antigenically distinct strains [12, 15, 16]. Following vaccination, vaccine-distinct variants might become dominant, requiring new vaccine formulations to be developed. One approach to overcome parasite diversity is to include multiple variants currently circulating in the global population, as for the Influenza vaccine which is reformulated annually [105]. Malaria differs from the Influenza however as malaria parasites have multiple lifecycle stages, hundreds of antigens that could be considered as vaccine candidates, and a lack of understanding of how parasite antigen diversity relates to host immune responses. With the availability of thousands of malaria parasite genomes [18], characterisation of antigen diversity on a global scale is highly feasible and a crucial step in contemporary malaria vaccine development. A meta-population genetic analysis of antigens would provide a catalogue of common variants to be included in vaccine formulations and provide a basis for predicting vaccine effectiveness in different populations.

Malaria antigen diversity has been largely overlooked with most subunit vaccines based on a single antigen variant, predominantly those of the reference strain 3D7. As a result, very few have shown significant protective efficacy in human clinical trials, and protection is mostly short-lived. Of the numerous vaccine candidates that have been tested in clinical trials, only RTS, S, has completed Phase-3 clinical trials. RTS,S is based on the C-terminal and NANP repeat region of the Circumsporozoite Protein (CSP) of the African strain 3D7 [12, 13]. The limited efficacy of this vaccine [14] has been attributed to polymorphism of the target antigen, PfCSP, suggesting a diversity-covering approach could be more effective [12,15, 68]. This is supported by a study in Africa where the one year cumulative vaccine efficacy against homologous strains was 50.3% compared to 33.4% against heterologous strains [5]. Furthermore, in a phase 2b trial in Papua New Guinean (PNG) children, the “Combination B” malaria vaccine (multiantigen vaccine, composed of MSP2, RESA and a fragment of MSP1) showed high protective efficacy against vaccine like strains and limited efficacy against vaccine dissimilar strains [16]. Of the vaccine trials that have been powered to measure allele specific outcomes (comparing infection rates of vaccine versus non-vaccine variants in vaccinees and controls), several have identified variant-specific efficacy (reviewed in [97]). Vaccine development would therefore benefit from a better understanding of the global diversity and distribution of antigen variants, to select common variants that could maximise vaccine efficacy.

The presence of intermediate variant frequencies for a given antigen in a parasite population indicates balancing selection, which is driven by protective immune responses [7–9]. Balancing selection can be measured using the *Tajima’s D* statistic using a ‘sliding window’ approach to identify hotspots of selection, along the length of a gene [10]. However, standard Tajima’s D analyses of linear gene sequences do not consider the spatial distribution of polymorphic residues (i.e. residues that are distant on the linear sequence may be proximal on the three-dimensional (3D) structure). Despite the availability of high quality whole genome sequence data from thousands of worldwide *P. falciparum* isolates, there has been no (meta)-population genetic analysis of malaria vaccine candidate antigens using this data, and only a few studies have mapped selection hotspots onto experimentally-determined or modelled 3D protein structures [9, 11]. In addition, only a few studies have analysed the diversity of existing or emerging *P. falciparum* vaccine candidate antigens and the majority have focused on few antigens and geographic areas.

In this study, we aimed to characterise the global genetic diversity of 23 emerging and established *P. falciparum* vaccine candidates from different life cycle stages and sub-cellular localizations, as described in the WHO Malaria Rainbow Table [23]. Antigens were selected based on function or inclusion in pre-clinical or clinical (human) vaccine development studies [2]. They include antigens that have well-defined three dimensional (3D) structures as well as disordered proteins that are not well characterised [17]. The analysis was based on *P. falciparum* whole genome sequences (WGS) from the Pf3K (version 5.1) global data resource [18] which comprises more than 2,600 (WGS) from 15 countries and 156 additional WGS from PNG [19] generated in collaboration with the MalariaGEN *P. falciparum* Community Project. These comprehensive population genetic analyses of malaria vaccine candidates will aid the development of more broadly effective vaccines.

## Results

### High haplotype diversity and balancing selection indicates antigens that are natural targets of host immunity

The final sequence dataset included 1079 to 1499 sequences per antigen and a mean of 107 sequences per country (n = 26 to 433) (Table 1, Table S1). Nigeria was removed from downstream analysis due to very low sample size (n = 4).

**Table 1:**
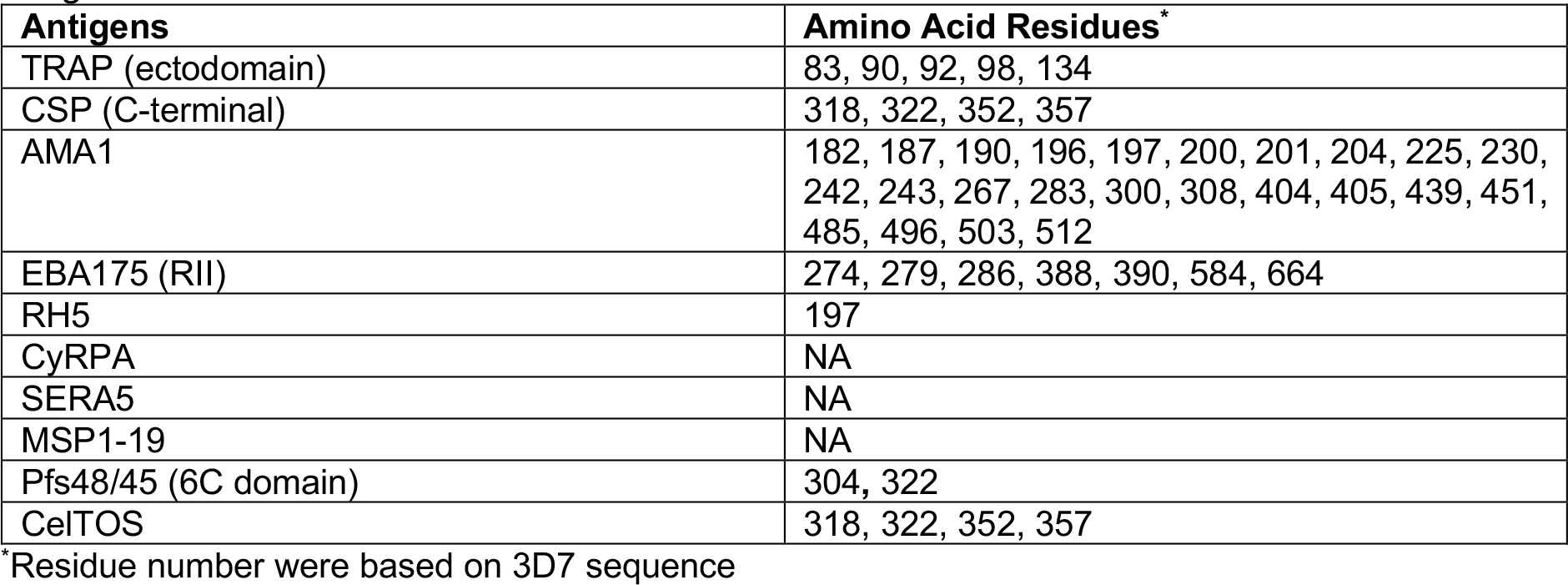
Vaccine Candidate Antigens. P. falciparum antigens and associated domains selected for the analysis were functionally important for malaria biology and key candidates of malaria vaccines trials^2^.

Most antigens showed high proportions of SNPs that were non-synonymous and regions of balancing selection [34] (Table S1, Fig 1). In addition, the antigen subdomains, *csp*-C-terminal, *eba175* -RII and *pfs48/45* -6C were as diverse as the respective full-length gene (Fig 1, Table S1). Different patterns of diversity across populations were observed for the antigens. For instance, *trap* had high haplotype diversity with low to moderate nucleotide diversity indicating that many haplotypes were the result of very rare polymorphisms (Fig 1, Table S1). Whereas *csp* had high haplotype diversity and high (but variable) nucleotide diversity, which might be related to differences in transmission intensity in different locations (Fig 1). In addition, *celtos* (a gametocyte antigen) also had high haplotype diversity but moderate to high (and variable) nucleotide diversity, which may indicate adaptation to mosquito vectors from different geographical regions (Fig 1, Table S1). Moderate haplotype diversity, but low nucleotide diversity for *rh5* indicates lower diversity among distributed *rh5* haplotypes (Fig 1, table S1).

**Figure 1.**
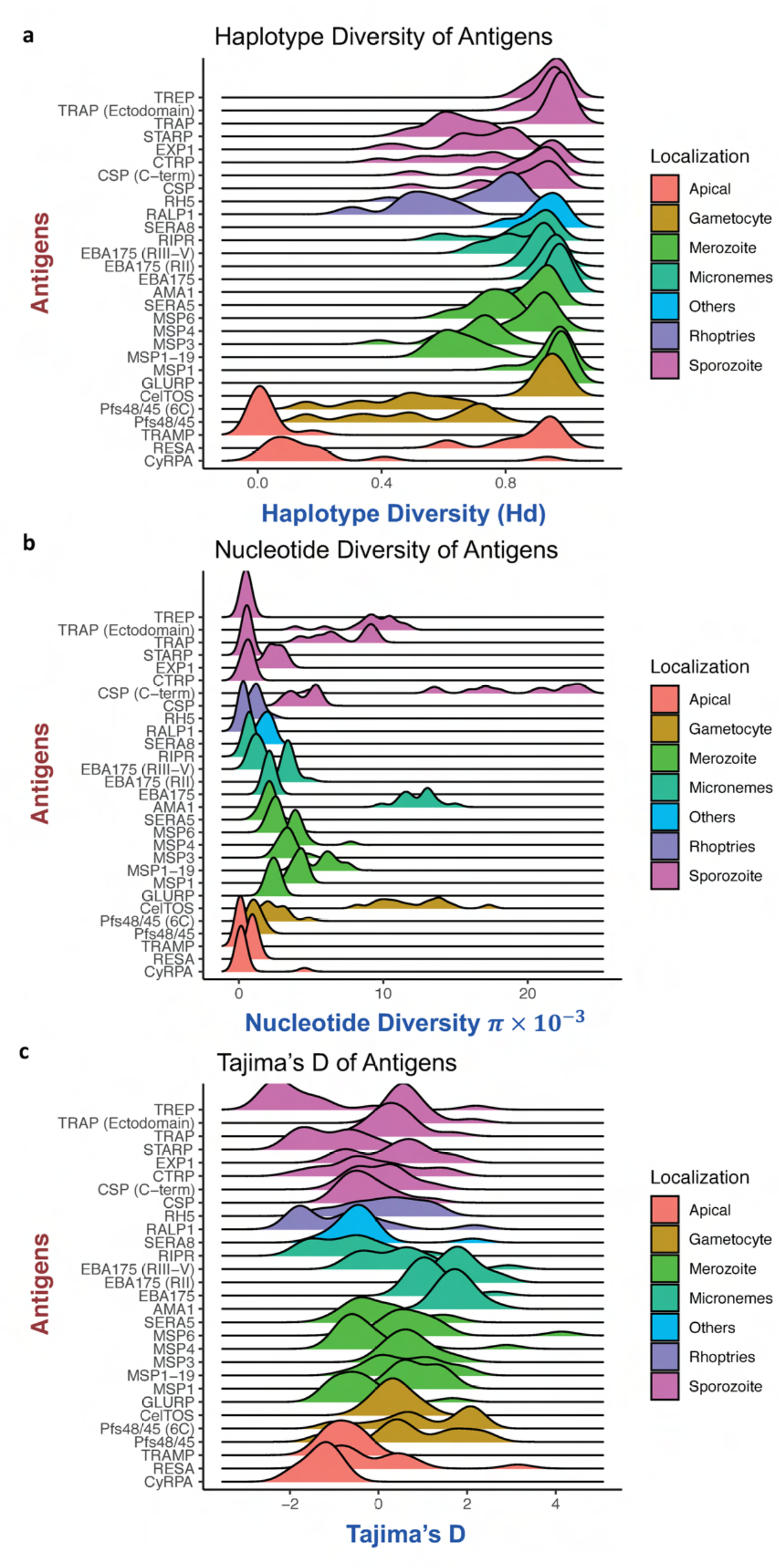
Distribution of haplotype diversity, nucleotide diversity, and Tajima’s D values amongst countries for each antigen. The ridgeline plots indicate the distribution of respective diversity matrices for each antigen from multiple geographic area or countries from MalariaGEN and PNG database. Tramp, and cyrpa were conserved across all populations. On the other hand, trap, ama1, eba175, and celtos were diverse and showed evidence of diversifying selection across all geographic area or countries.

Genetic diversity and evidence of diversifying selection on the antigens was not related to subcellular localization [35] (Fig 1). However, the diversity of some antigens (e.g. CSP) varies with geographic origin indicating that transmission intensity or other location-specific factors may play a role as previously shown in Barry *et al*. 2009 [7]. The use of full-length or full-domain *Tajima’s D* values can mask variable patterns within a gene when there are discrete stretches of positive *D* values linked by regions with neutral or highly negative D values [32]. Further analyses were therefore performed on these antigens for full-length or their functionally important domains.

Relative solvent accessibility (RSA) gives an indication of the regions of a protein that are exposed to the extracellular environment, and thus may be targeted by immune responses. The RSA of all 23 antigens was compared for 792 polymorphic and 14,789 conserved residues. A higher RSA score suggests presence of a particular amino acid on the surface and thus able to interact with the host environment. Overall, when combining all antigens, most of the polymorphic residues had significantly higher mean RSA scores of 0.46 than conserved residues with a mean RSA score of 0.38 (p value < 2.0 * 10^-16^, t-test) (Fig 2). When individual antigens were analysed, only AMA1, CSP, EBA175, MSP1, MSP4, Pfs48/45, SERA8, and TRAP had significantly higher RSA scores than that of their residues without underlying polymorphisms (p < 0.05, t-test).

**Figure 2.**
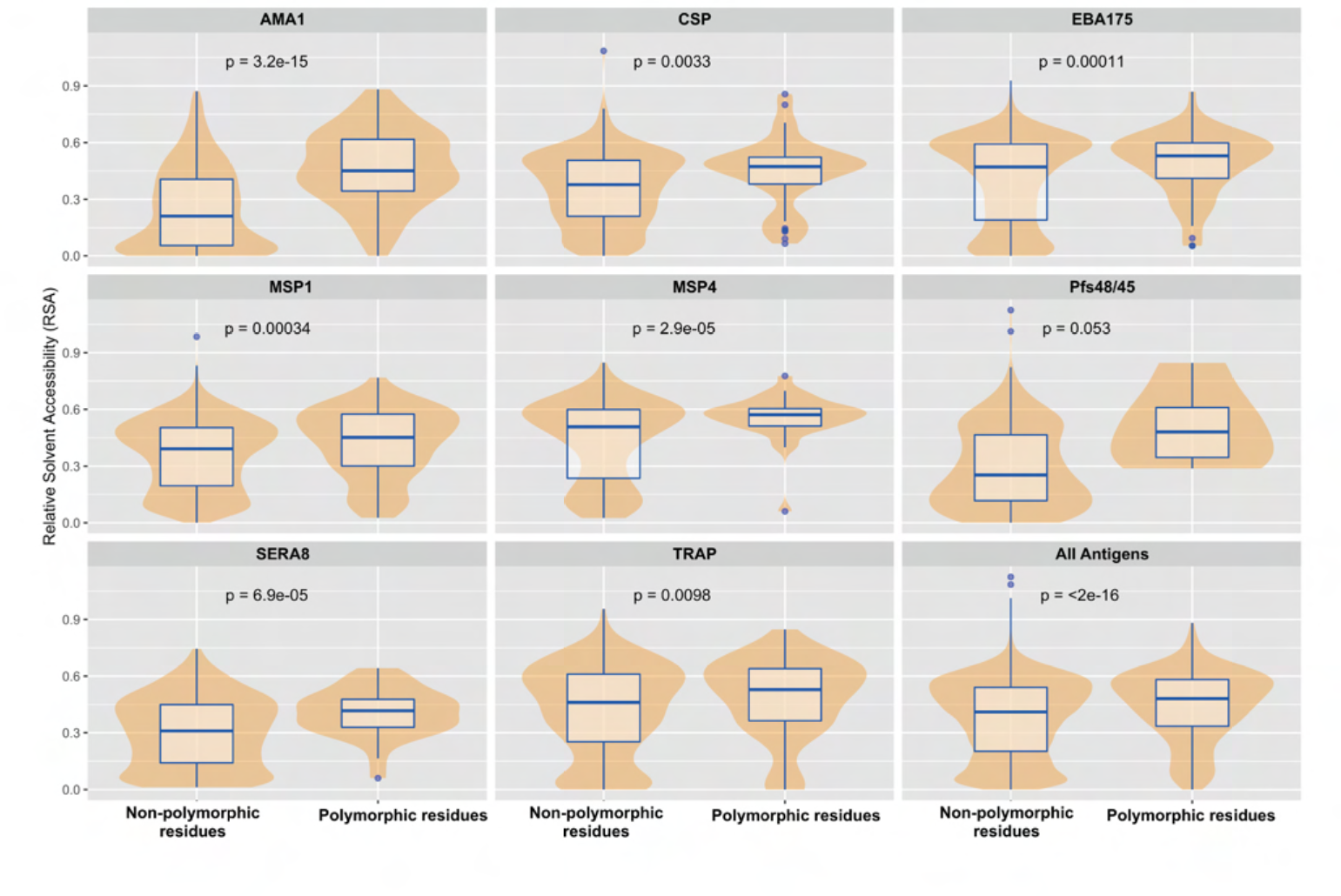
Relative solvent accessibility of polymorphic versus conserved residues. Relative solvent accessibility (RSA) was calculated for residues with and without identified polymorphisms from global dataset for all antigens or each antigen included in our analyses. RSA was calculated using neural network based NetSurfP1.1 program or DSSP program respectively based on the presence of known PDB or homology-modelled structures [29, 103]. Polymorphic residues from more than 1000 sequences regardless of minor allele frequency (MAF) were included in the analysis. Box and whisker plots show the median (blue line), and interquartile range (blue box) of RSA values for each residue from respective group. The violin plot (which uses Kernel Density Estimation to compute an empirical probability distribution) shows a smooth distribution of RSA values for most of the calculated group. RSA scores for individual antigens as well as for combination of all antigens are calculated. Only significant p-values are shown.

### Polymorphic residues are predominantly found at functionally important interfaces

Shannon Entropy is a measure of the variability of individual amino acid positions within a protein. We calculated the normalized Shannon Entropy using all available sequences for each antigen with an available 3D structure (Table 2) to investigate the potential functional impact of site-specific diversity. Except for CyRPA, SERA5 and MSP1-19, the normalized Shannon Entropy scores were high (>0.2) among the residues situated at host-receptor binding interface, within previously described immunological epitopes or under balancing selection (Fig 3, table 2). For instance, residues situated at the surface exposed c1L loop of AMA1, and residues situated at the dimerization interface of EBA175 RII had high normalized Shannon entropy scores. Parasites may adapt the amino acids more frequently at functionally important interfaces to escape from the host immune system.

**Figure 3.**
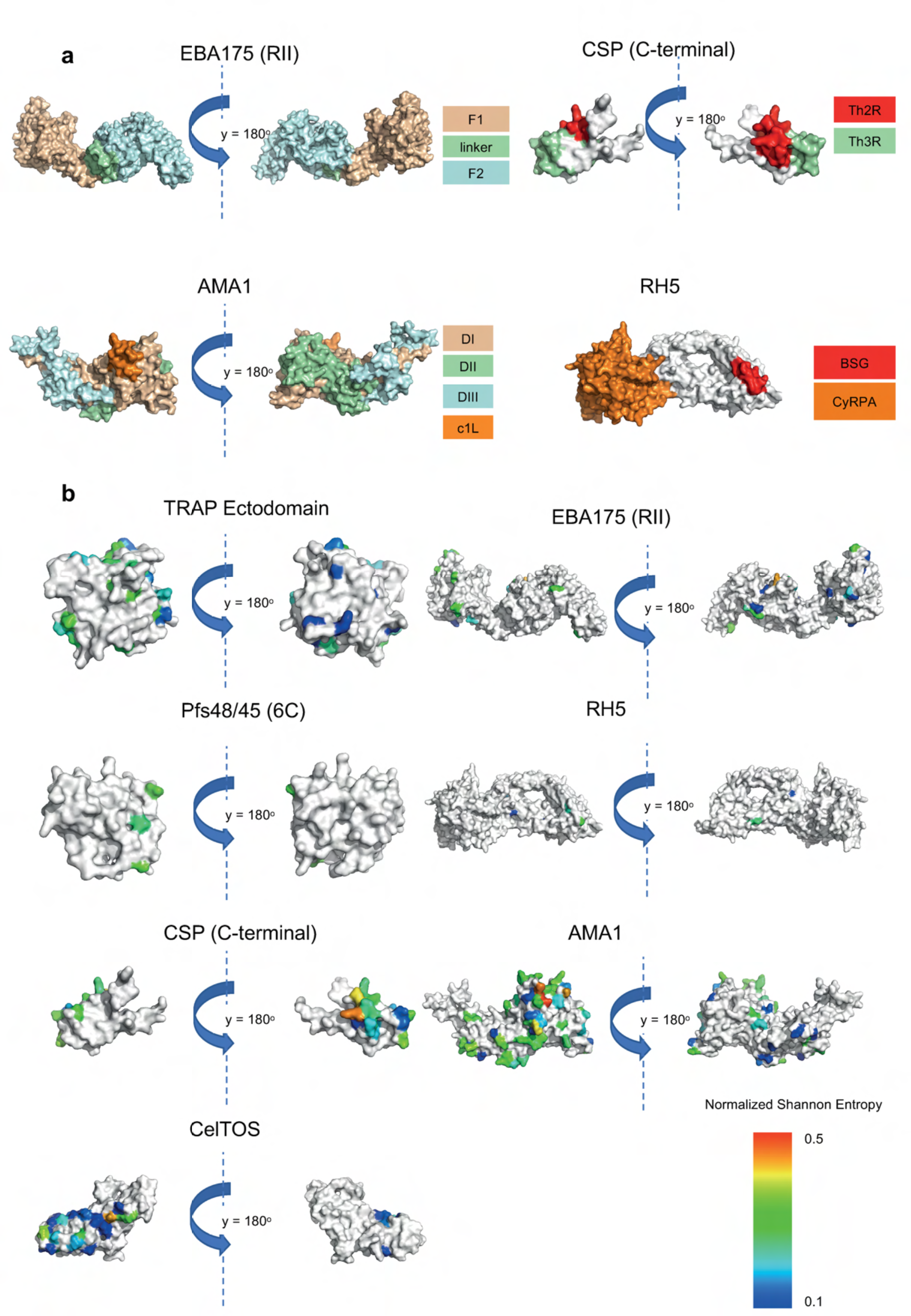
Mutations at functionally important interfaces. **(a)** Available domain and epitope information for AMA1, EBA175 (RII), RH5, and CSP (C-terminal). **(b)** Site specific diversity measure for CelTOS, Pfs48/45 (6C domain), TRAP (ectodomain), CSP (C-terminal), AMA1, EBA175 (RII) and RH5. Normalized Shannon Entropy was calculated per residue for these antigens (unavailable residues were coloured in white). Higher entropy values indicate higher diversity across all populations for a particular residue. Residues from the CSP Th2R epitope and AMA1 C1L loop have the highest entropy values. Very low entropy values across all populations were observed for SERA5, and CyRPA (not shown).

**Table 2.**
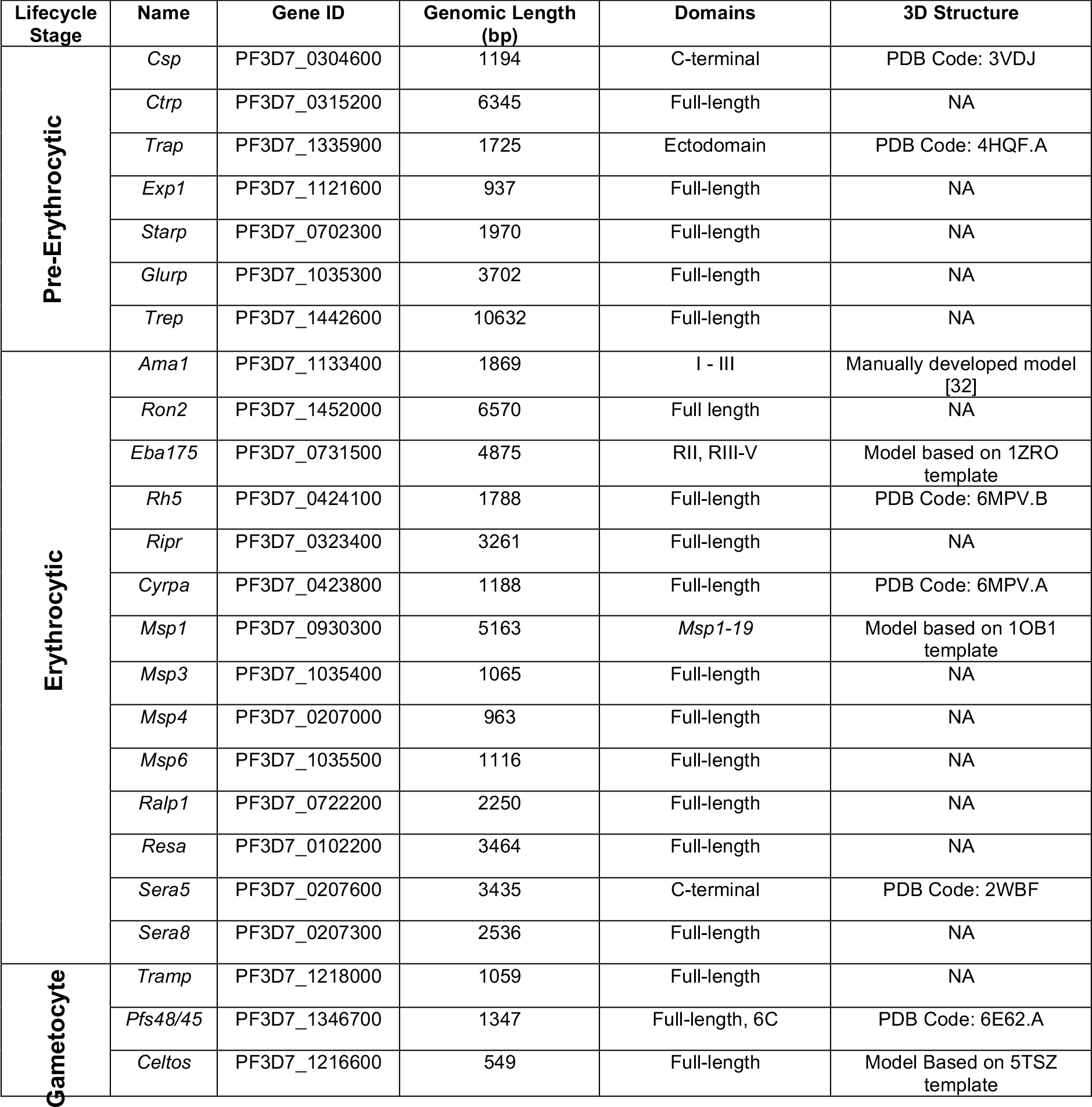
Residues with Shannon Entropy Scores greater than 0.20 for each calculated antigen.

### Evidence of balancing selection at functionally important interfaces for a subset of key invasion ligands

An ideal malaria vaccine should be immunogenic and effective against naturally circulating strains from worldwide populations, but it is not feasible to include all haplotypes for each antigen we have described above. This is especially the case for highly diverse antigens such as CSP, TRAP, AMA1, EBA175, MSP1 (table S1). Therefore, for each antigen, it is important to identify the most immunologically relevant polymorphisms and gene regions, that are targets of host immunity and could influence vaccine efficacy. Typically, the *Tajima’s D* statistic has been calculated as a single metric encompassing the entire gene or domain of interest, or more recently, using a sliding window approach to identify hotspots of balancing selection along the length of a gene. Functional antibodies are critical for protective immunity from naturally acquired infection [12,71–74] however, more than 90% of functional antibody epitopes are discontinuous (non-linear) epitopes [75, 76]. Thus, distant gene segments may be brought into proximity in three-dimensional (3D) protein structures. Consideration of structural features in 3D space are thus important, especially for antigens with highly surface exposed polymorphic residues such as AMA1, CSP, EBA175, and TRAP. Previous studies have suggested that polymorphic residues located on the surface of protein evolved to escape host immune responses [9,32,36]. We therefore examined selective pressure over the 3D protein structures of vaccine candidate antigens where possible.

A spatially derived approach to *Tajima’s D* (D*) was applied to available 3D structures (predicted or experimentally defined) for a subset of parasite populations covering all major endemic regions, including Southeast Asian, Africa, and Oceania (PNG). Structures included well-studied antigens like CSP (C-terminal), TRAP (ectodomain), AMA1, EBA175 (RII), MSP1, RH5, CyRPA, SERA5, CelTOS, and MSP1-19. We observed moderate to high D* scores (1.0 - 2.0), high scores (2.1 - 3.0), and extremely high scores (> 3.1) within most of these antigens, which is an indication of balancing selection focused on specific regions of the protein. Nearly neutral *Tajima’s D* values were found throughout the 3D structure of CyRPA, and SERA5 (C-terminal) for all parasite populations (Fig S4). In general, we found a dichotomous pattern of balancing selection on some antigens for countries in the Asia-Pacific versus African populations.

The TRAP ectodomain (PDB Code: 4HQF.A) which is comprised of tandem von Willebrand factor A (VWA) and thrombospondin type I repeat (TSR) domains (AA residues: E41 – K240, 3D7 sequence) were included in our analyses [37]. In all populations, the suspected heparin binding interface of TRAP [38] has moderately higher *Tajima*’s *D\** scores than the opposite interface of the protein, which acts like the silent face of TRAP (ectodomain) (Fig 4). Hence, residues Y89 – I100, and I115 – D146 showed moderate spatially derived *D*\* scores (1.0 - 1.2) in all observed countries. Residues S123, T124, and N125 form a minor protrusion on the surface of TRAP, and residues R130, R141, K142 are thought to be mediated in heparin binding according to previous study [38]. However, within the active face of TRAP, the intensity of balancing selection (*D\** scores) appears to be geographically variable. However, minimal nucleotide diversity variation was observed amongst geographical regions (Fig 1).

**Figure 4.**
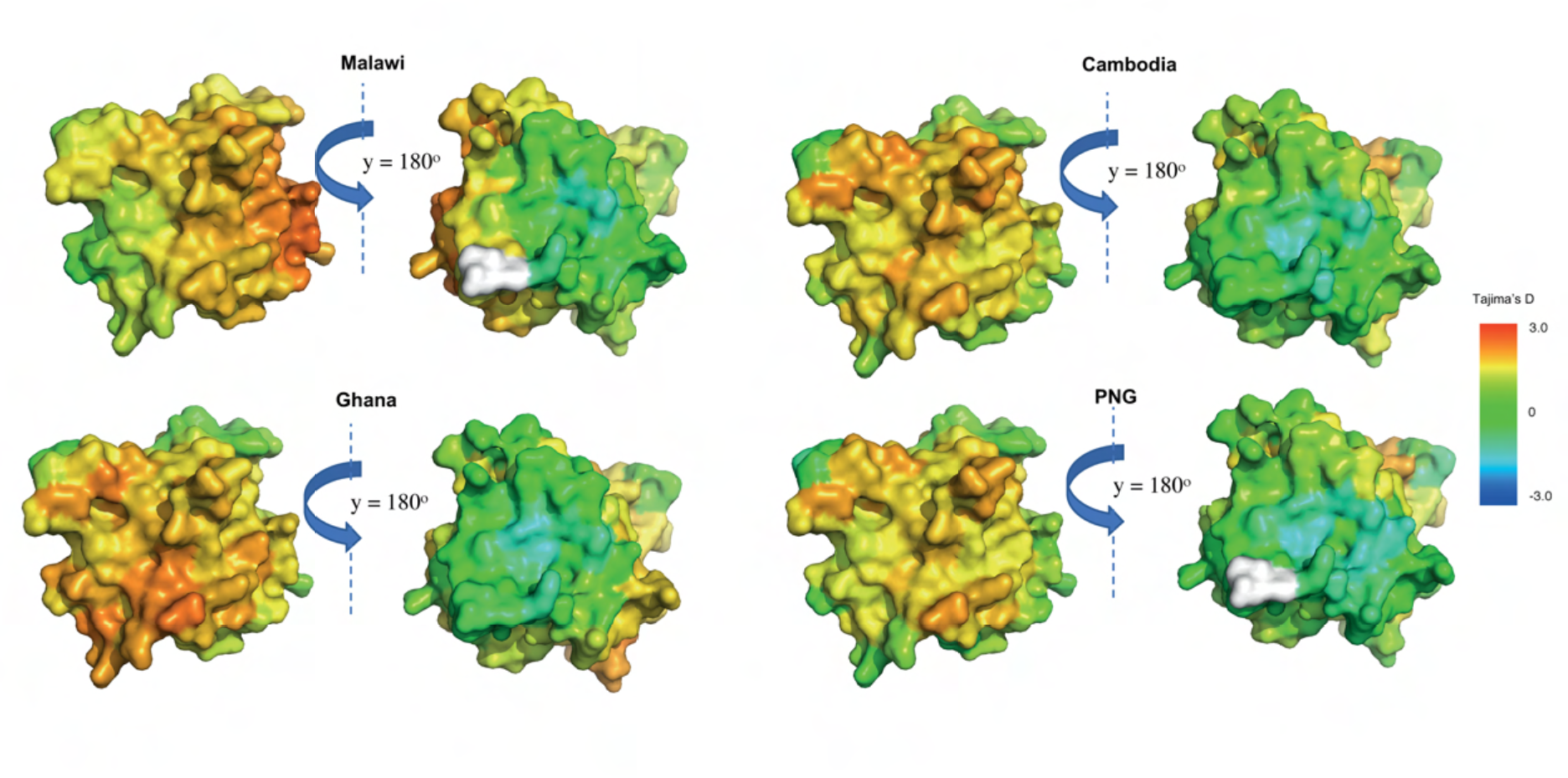
Evidence of balancing selection pressure at functionally active interface for TRAP (Ectodomain). Spatially derived Tajima’s D (D*) score calculation for TRAP (Ectodomain) with incorporation of protein structural information using a 15Å window. TRAP (ectodomain) (PDB Code: 4HQF.A) was used. The structure was coloured according to D* scores mapped to each residue, and undefined D* are shown in white. Sample sizes: Malawi (n = 133), Ghana (n = 238), Cambodia (n = 430), and PNG (n = 112).

High *D\** scores (2.0 – 2.4) were found in all populations at the C1-L cluster [39, 40] of AMA1 Domain I (AA residue: T194 – D212, 3D7 sequence), known to be associated with immune escape [9, 39, 40, 45] (Fig 5). Most of these residues were identified as discontinuous epitopes by the IEDB epitope prediction resource [41]. As expected, the entire AMA1 interface with hydrophobic binding cleft and RON2 interacting sites shows high balancing selection [42]. Similar to previous spatially derived analysis [9], the region on the border of DII and DIII domains (AA residues: P303 – F312, S432 – Y446, I479 – K508, 3D7 sequence) appears to have the highest *D\** scores (> 3) in all populations. This region was previously predicted as the surface exposed face of DII/DIII and suggests that these residues might be the targets of protective host immune responses within AMA1. A monoclonal antibody (1E10) against AMA1 DIII functions synergistically with antibodies against distant parts of AMA1 to inhibit merozoite growth [43]. This indicates that DIII of AMA1 plays a significant role as a target of functional antibody responses against *P. falciparum* in the context of natural infections. Consistent with previous findings [9,32,44], the spatially derived *Tajima’s D* values are unevenly distributed with high *D\** values mostly exclusive to one face of the AMA1 molecule, and close to zero on the opposing face of AMA1, which has been previously been described as the “silent face” [99, 100] except in PNG where moderately high *D\** scores (1.9) were found at AA residues V524 – Y532. This is in line with the previous hypothesis that this region of AMA1 has minimal exposure to the immune system [44, 45]. The high level of spatial Tajima’s *D\** and nucleotide diversity was similar amongst populations for AMA1 suggesting that this antigen experiences similar selective pressures across different populations (Fig 5, and S5).

**Figure 5.**
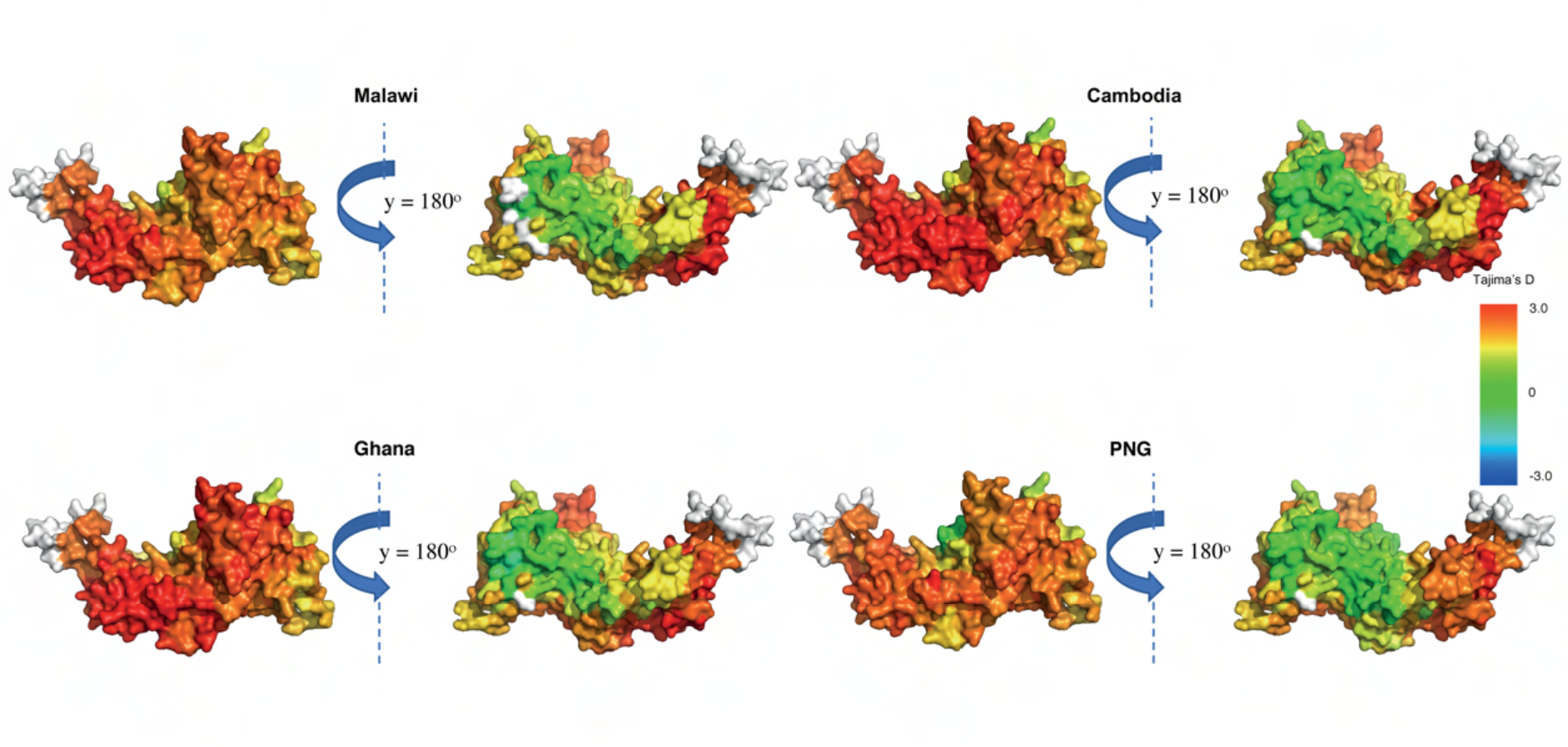
Geographically conserved selection pressure for AMA1. Tajima’s D (D*) calculations for AMA1 with incorporation of protein structural information using 15 Å window. The manually modelled structure of AMA1 was used based on published results [32]. The structure was coloured according to D* scores mapped to each residue with undefined D* scores were shown in white. Sample sizes: Malawi (n = 139), Ghana (n = 243), Cambodia (n = 433), and PNG (n = 112).

*Ripr* was analysed in the context of the linear nucleotide sequence due to the poor predicted structure. The N-terminal region of RIPR contains 2 EGF-like domains, while the C-terminal region contains 8 EGF-like domains [46]. We found some degree of balancing selection (moderately high *D* values = 1.5) and high nucleotide diversity proximal to C-terminal EGF 7-10 region, located away from the CyRPA:RIPR interface [47] (nucleotide residues based on coding region 2900-2980, 3D7 sequence) of *ripr* at most of the populations (Fig 6). These results were aligned with the anticipated RIPR-specific monoclonal antibody binding sites identified in a previous study [47]. This suggests the C-terminal region of RIPR is a target of host immunity in most populations.

**Figure 6.**
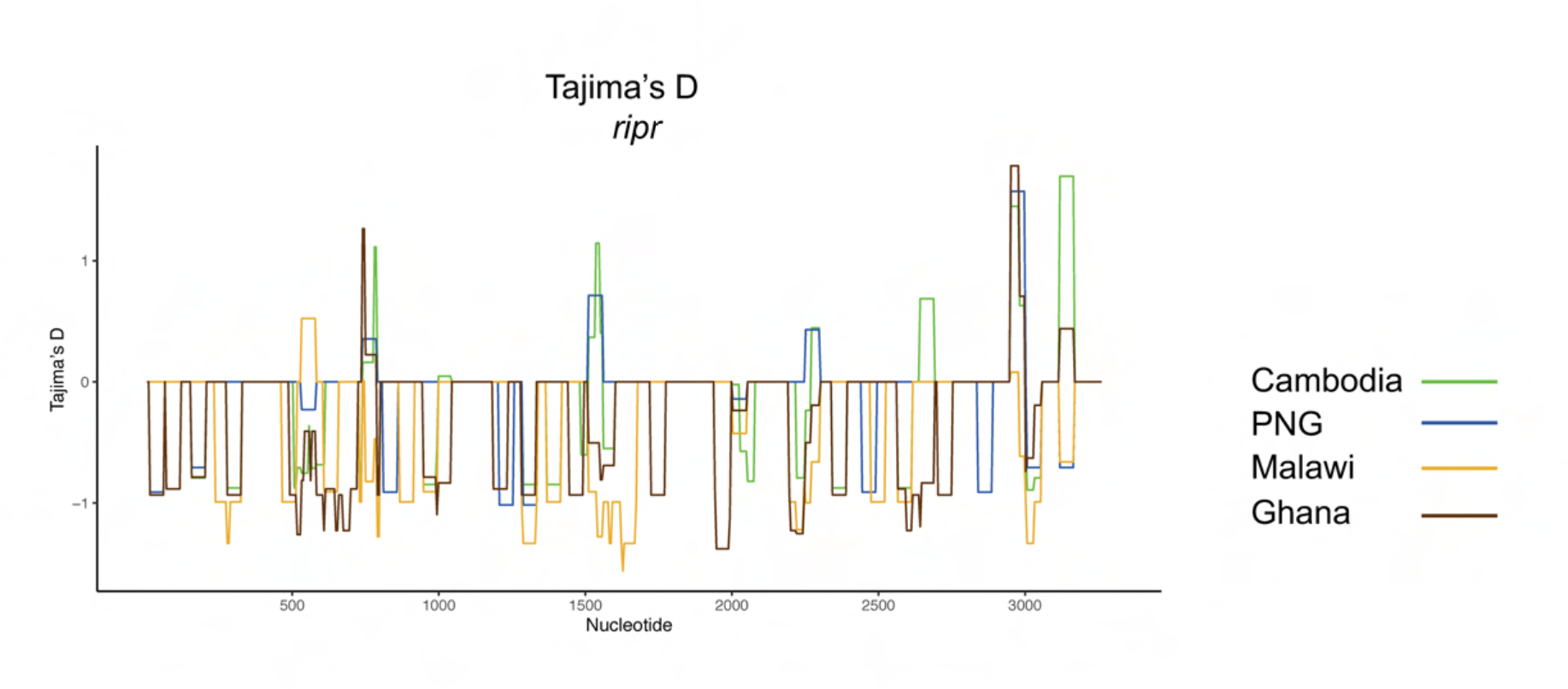
Polymorphism and evidence of selection for ripr. Tajima’s D statistic calculated for disordered regions of RIPR in samples from Cambodia, PNG, Malawi, and Ghana. Tajima’s D is calculated with a sliding window approach (a window size of 50 bp and a step size of 5 bp). Nucleotide positions based on coding region are shown in the x-axis. Sample size for each respective population are as follows: Malawi (n = 137), Ghana (n = 246), Cambodia (n = 428), and PNG (n =111).

### Balancing selection hotspots vary geographically for EBA175 (RII), RH5, CSP, CelTOS, and MSP1-19

While the crystal structure of EBA-175-RII has been solved [48], we used a 3D7-based *ModPipe* homology structure for analysis, as this model includes a number of functionally important residues that were not resolved in the experimental structure [8]. For *D\** analysis on EBA-175-RII, a large region of the F1 domain which predominantly consisted of AA residues E226 - L294, I312 – K324, and W377 – I400 was under balancing selection (i.e. high *D\** scores of 2.8 – 3.0) in all parasite populations. This cysteine-rich region is also involved in the dimerization interface formation (helix linker and disulphide bridges)[48] between two molecules of EBA-175-RII as it binds to human receptor glycophorin A. During dimerization, this region also makes contact with another cysteine rich F2 domain of the other dimer pair in a ‘handshake’ interaction [48]. However, slight variations between African and Asia-Pacific populations can also be found within the site from F2 domains, which are predominantly comprised of AA residues C476–C488 and Y710 – F722 where high *D\** scores (1.8 – 2.0) was observed within most Asian-Pacific countries, but not African countries (*D\** scores < 0.4) (Fig 7). The F2 domain makes most of the contact with glycan [33] and residues C476–C488 are a part of F2 β-finger domain that is also a target of inhibitory antibodies, R215 and R217 [49, 50]. The linker region was found to be conserved within all populations (Fig 7). However, the limited nucleotide diversity was observed amongst geographical regions (Fig not shown).

**Figure 7.**
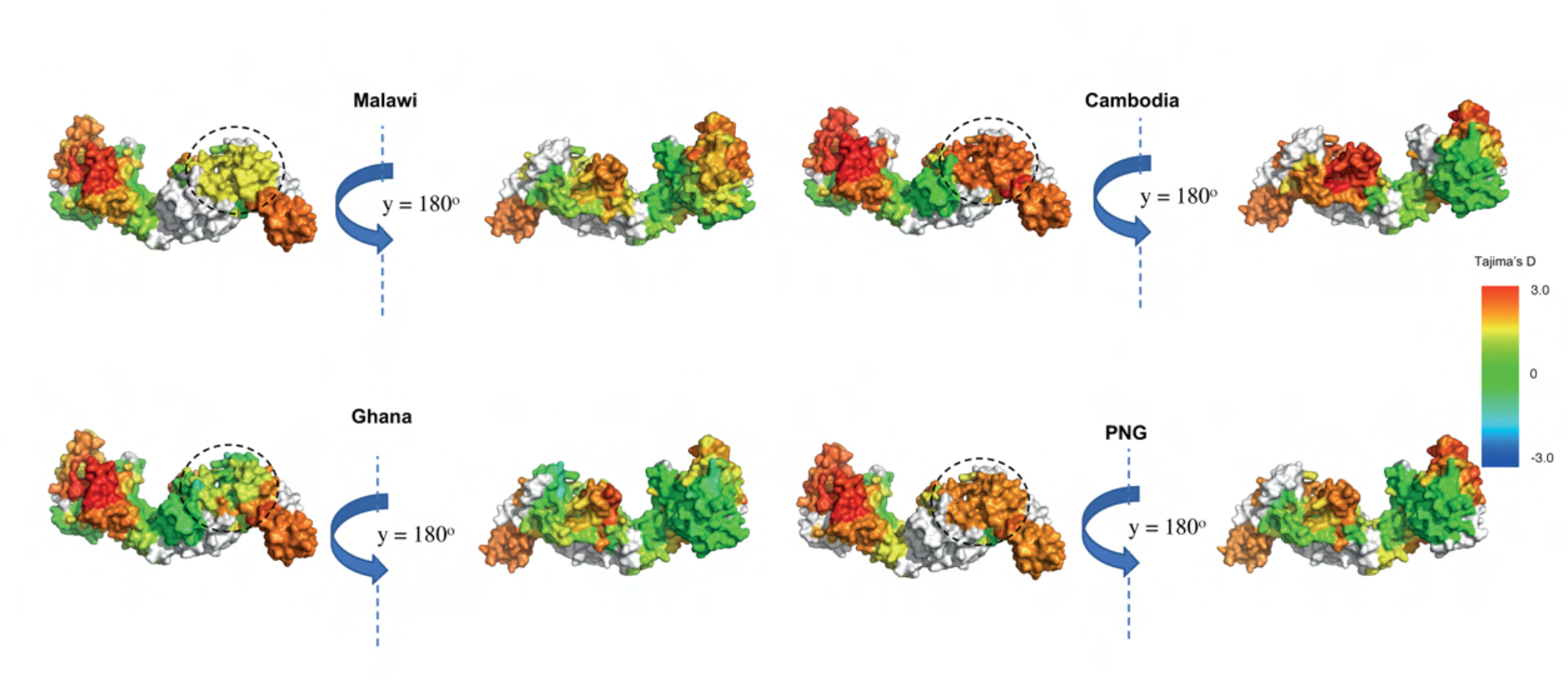
Geographically variable selection pressure on EBA175-RII. Spatially derived Tajima’s D (D*) for EBA175 with incorporation of protein structural information using 15 Å window. 3D7-based ModPipe model of EBA175 (RII) based on 1ZRO template was used. Structure was coloured according to D* scores mapped to each residue with undefined D* were shown in white. The highlighted region (in circle) shows different D* scores between Asian-Pacific and African countries. Sample sizes: Malawi (n = 136), Ghana (n = 237), Cambodia (n = 432), and PNG (n = 112).

The cryo-EM structure of full length RH5 (PDB Code: 6MPV, chain B) was used in our analysis [51]. Evidence of balancing selection was limited within African countries parasite populations based on the spatially derived *Tajima’s D\** scores. In contrast, within Asia-Pacific countries, moderate to high *D\** scores (1.5 - 2.0) were consistently observed at the Basigin (BSG or CD147) binding sites and appear to be under balancing selection (AA residues: S189 – D207, 3D7 sequence). Additionally, the residues near CyRPA binding sites which predominantly consist of residues I386 – F421 have moderate evidence of balancing selection (*D\** score of 1.5) within PNG population. This suggests the BSG binding site may be a key target of host immunity and aligns with previous findings from Alanine *et al.* (2019) [52]. Non-synonymous AA polymorphisms at residues Y147 and H148 (nucleotide positions 439, and 442, 3D7 sequence) were under balancing selection in most Asian populations but appear to be under adaptative selection in some African populations according to the linear sliding window analysis with *Tajima*’s *D* values around 1.7 (Fig S3). These residues did not have PDB coordinates (physical proximity to N-terminal disordered region), and therefore cannot be detected via spatially derived 3D analyses. Nucleotide diversity of RH5 was slightly higher in Asian-Pacific populations than African populations (Fig S3).

We observed limited evidence of balancing selection (near neutral) within the thrombospondin receptor domain of CSP for Asia-Pacific populations (Cambodia and PNG). However, moderately high *D\** scores (1.2 – 1.3) were observed at Th2R residues (AA residues E310 – L327, 3D7 sequence) at Malawi and Ghana populations which suggests some evidence of balancing selection (Fig 9). Similarly, nucleotide diversity of some of the residues comprising thrombospondin receptor domain of CSP were relatively low in Asia-Pacific populations compared to African populations (Fig S7). A moderately high to high degree of balancing selection (*D\** scores of 1.0 and 2.9) for Th3R residues (AA residues G341 – I364) was also observed within African populations (Fig 9).

**Figure 8.**
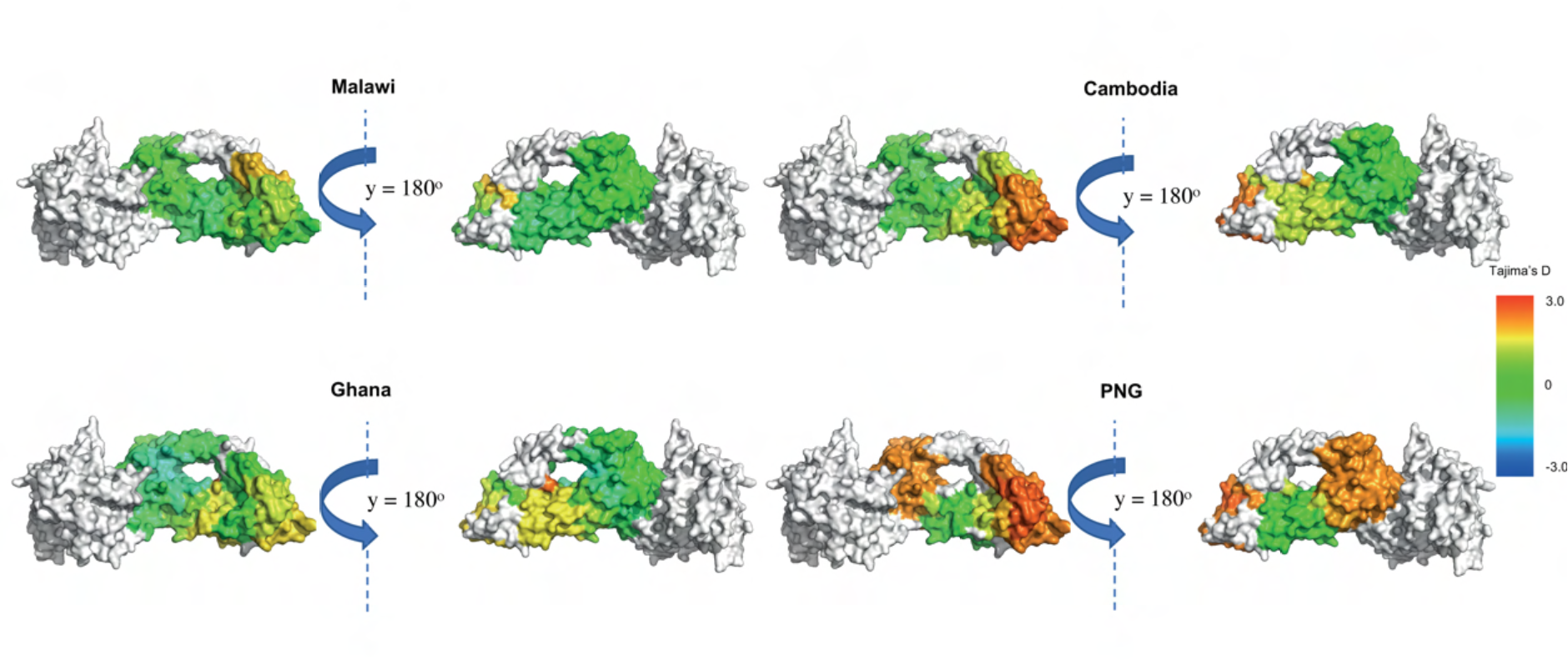
Geographically variable selection pressure for RH5. Spatially derived Tajima’s D (D*) calculation for countries from Asian-Pacific and Africa for RH5 with incorporation of protein structural information using 15Å window. Cryo-EM structure of RH5-CyRPA complex (PDB code: 6MPV.B) was used. Structure was coloured according to D* scores mapped to each residue with undefined D* and CyRPA were shown in white. Sample sizes: Malawi (n = 142), Ghana (n = 249), Cambodia (n = 433), and PNG (n = 112).

**Figure 9.**
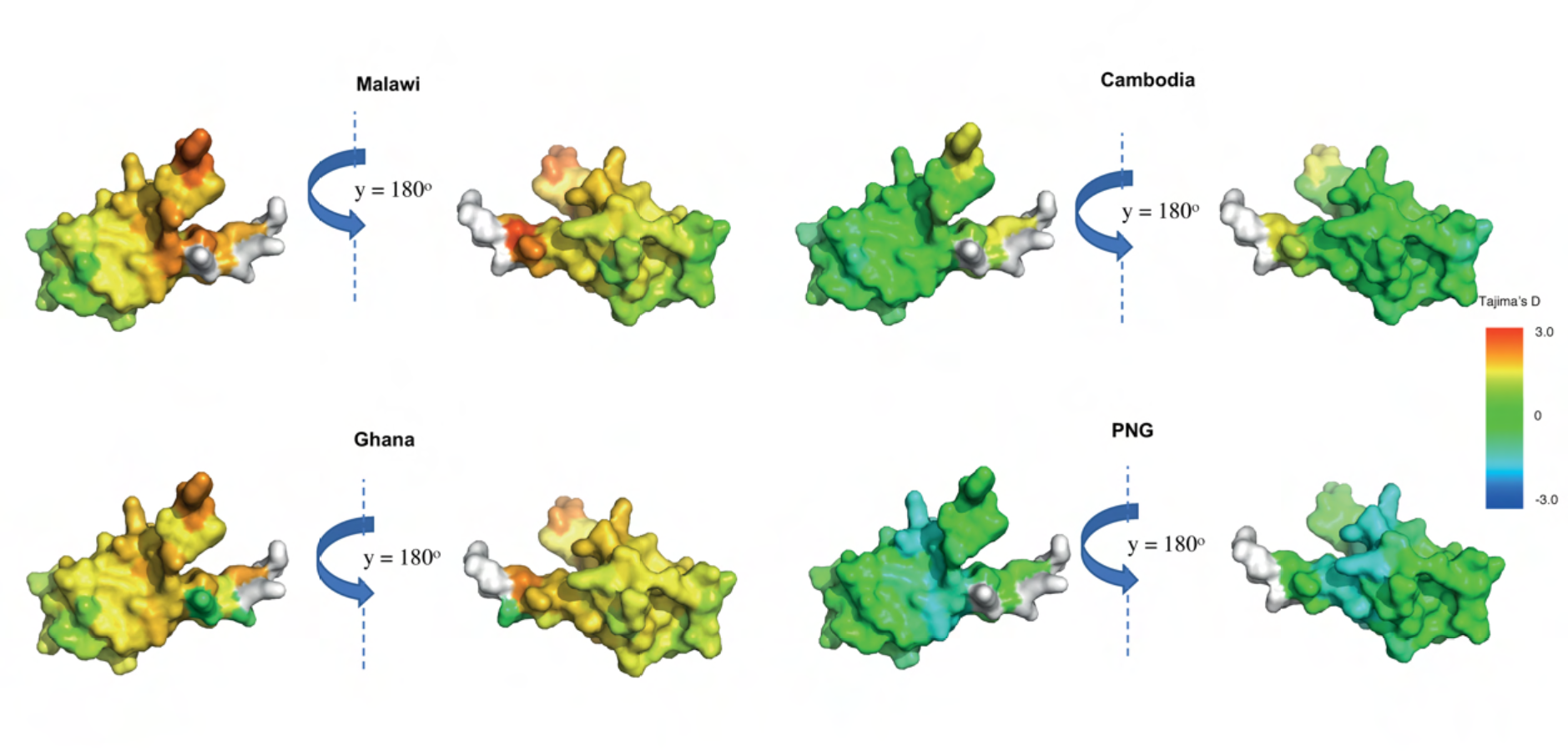
Geographically variable selection pressure for CSP (C-term) Tajima’s D (D*) calculations for populations from Asia-Pacific and African regions for CSP (C-terminal) with incorporation of protein structural information using 15 Å window. The crystal structure of the **t**hrombo**s**pondin **r**eceptor (TSR) domain of CSP [53] (PDB code: 3VDJ, chain A, AA residues: Y306 - H376, 3D7 sequence), which consists of Th2R and Th3R (T-cell epitopes) was used [54]. Structure was coloured according to D* scores mapped to each residue with undefined D* were shown in white. Sample sizes: Malawi (n = 135), Ghana (n = 223), Cambodia (n = 431), and PNG (n = 111).

Hotspots of balancing selection were found within N-terminal AA residues F87 and D93 – S104 (3D7 sequence) of CelTOS for most of the populations with moderately high to high spatially derived *Tajima’s D*\* scores (1.0 – 2.2) (Fig 10). Within the Malawi population, we found high *D\** scores (1.8 – 2.0) at AA resides D131 – I138 (3D7 sequence), but limited balancing selection was found for CelTOS within PNG (*D\** scores < 0.4) (Fig 10). Nucleotide diversity was also relatively low in PNG populations compared to others (Fig S7). However, the presence and distribution of *D\** values were variable amongst geographic locations (Fig 10). Limited functional information was available for CelTOS therefore the significance of these findings is not able to be postulated.

**Figure 10.**
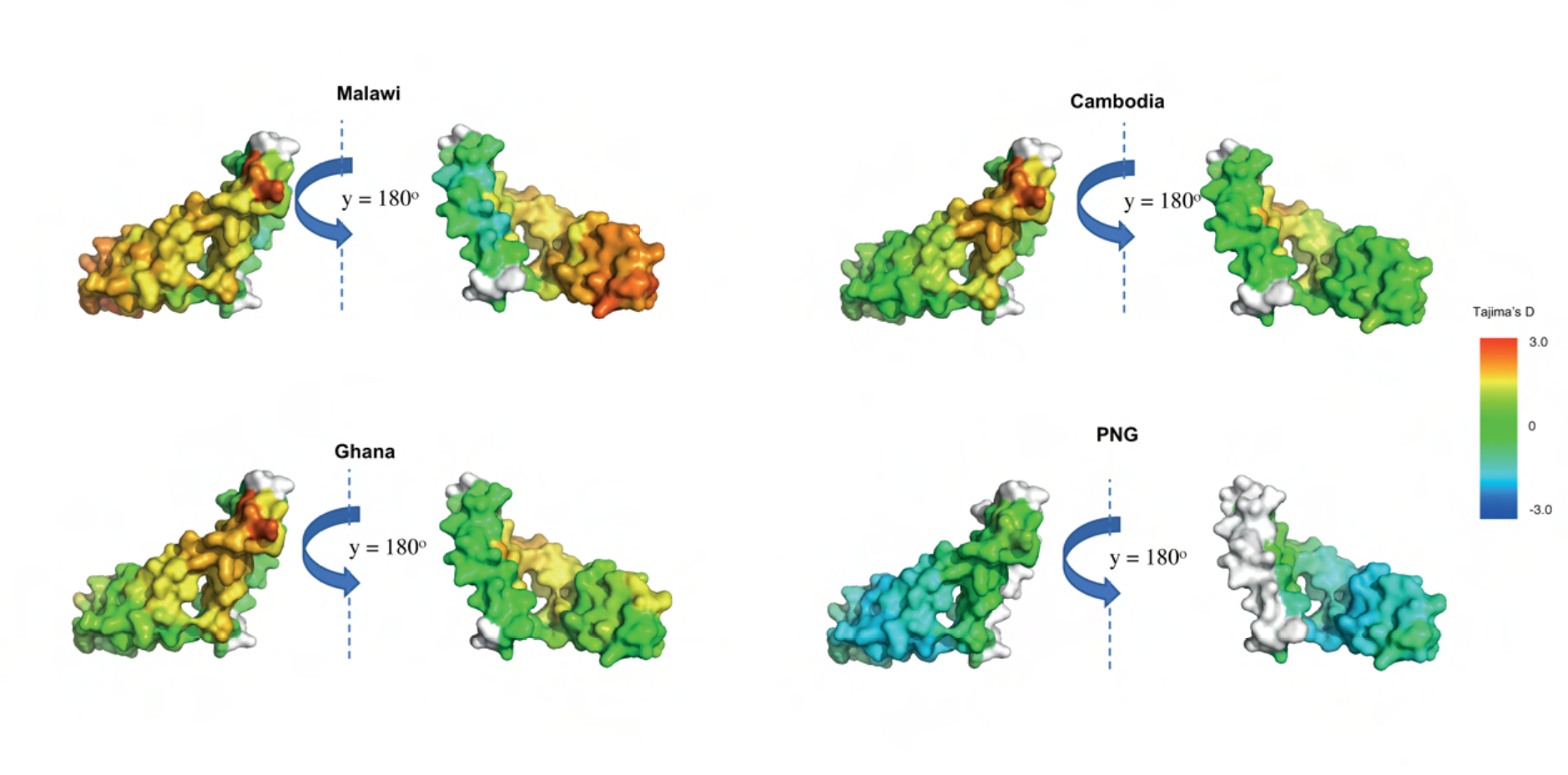
Geographically variable selection pressure for CelTOS. Tajima’s D (D*) calculations for populations from Asia-Pacific and African regions for CelTOS with incorporation of protein structural information using 15 Å window. Structure was coloured according to D* scores mapped to each residue with undefined D* were shown in white. 3D7-based ModPipe model of the P. vivax CelTOS based on 5TSZ template was used. Sample sizes: Malawi (n = 142), Ghana (n = 245), Cambodia (n = 433), and PNG (n = 112).

Moderately high *D\** scores (1.0 – 2.0) are found within residues Q1612 – F1625, and E1632 – C1647 from African populations (Malawi and Ghana), but not in Asian-Pacific populations. Most of these residues were a part of conformational epitopes and involved in interaction with potent monoclonal antibody (G17.12) [95] and inter-domain interactions. In addition, nucleotide diversity of these residues is relatively high in African populations compared to Asian-Pacific parasite populations (Fig S8). However, different part of MSP1-19 with residues P1651 – C1682 were under balancing selection in most of the Asian-Pacific populations (only shown here for Cambodia and PNG populations) (Fig 11).

**Figure 11.**
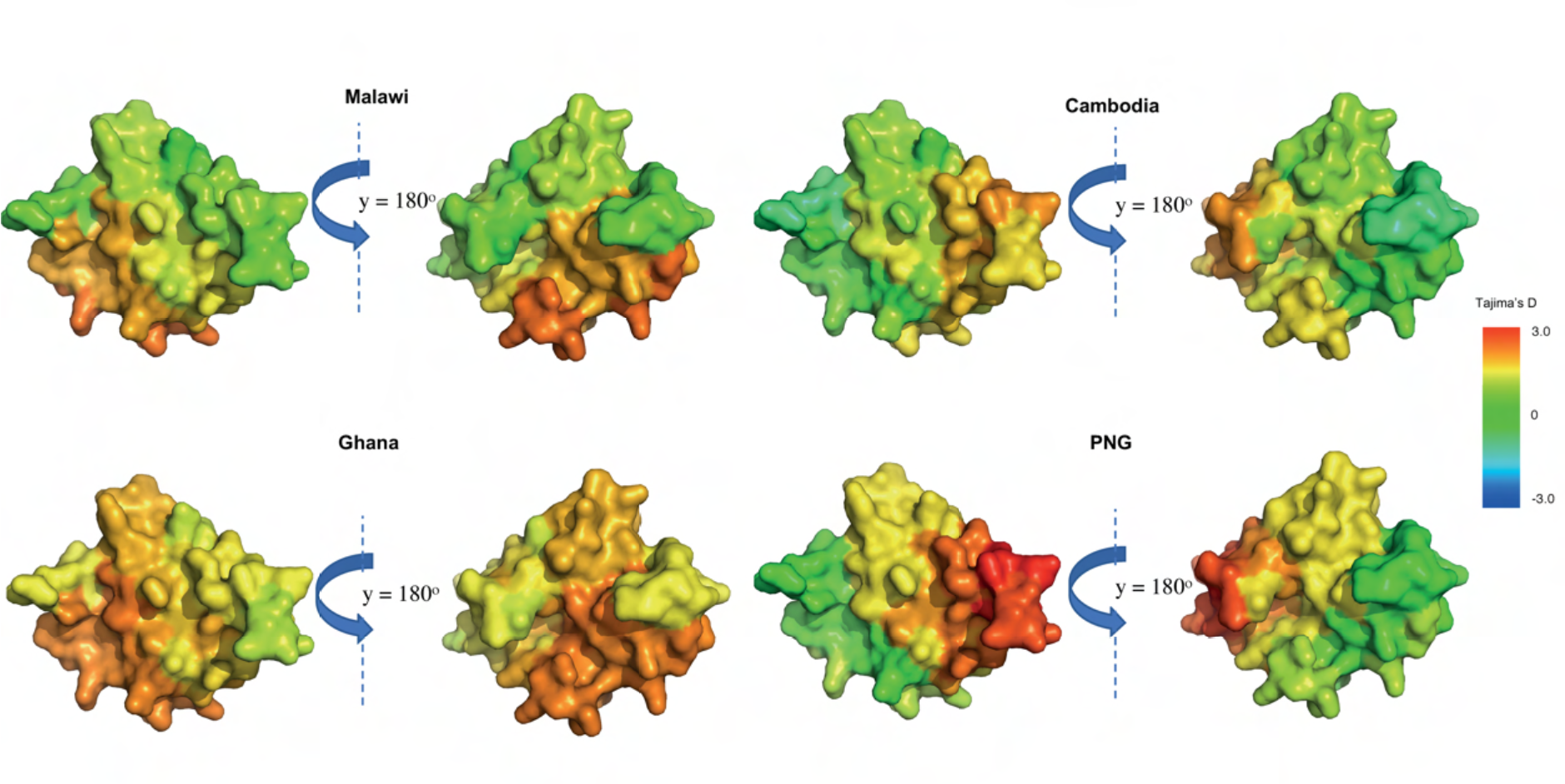
Geographically variable selection pressure for MSP1_19_. Tajima’s D (D*) calculation for geographic area or countries from Asian-Pacific and African regions for MSP1_19_ with incorporation of protein structural information using 15 Å window. The structured region of MSP1 (MSP1-19, AA residue: N1607 - S1699) based on ModPipe homology model using template (PDB code: 1OB1) was used [95]. The structure was coloured according to D* scores mapped to each residue with undefined D* were shown in grey. Sample sizes: Malawi (n = 101), Ghana (n = 183), Cambodia (n = 270), and PNG (n = 72).

### Intrinsically disordered proteins may be targets of immune selection

Intrinsically disordered proteins (IDP) and intrinsically disordered regions (IDR) are a newly recognised class of proteins, which lack secondary and tertiary rigid structure (under physiological conditions) but possess active roles in many biological processes [56–61]. Recent predictions have shown an abundance of IDPs within the *Plasmodium* proteome including several vaccine candidate antigens [30]. This is not surprising given that disordered regions create larger intermolecular interfaces, which increases the chances of interaction with potential binding partners, even without tight binding, providing flexibility for binding diverse ligands and other proteins including functional antibodies [84]. IDPs can have a diverse range of functions, although our understanding of the role of IDPs within biological systems is still incomplete [58, 101]. Due to the lack of well-defined three-dimensional structure for antigens with extensive disordered regions, we calculated linear *Tajima’s D* values to determine balancing selection. Highly disordered proteins were defined as those with more than 50% residues with disorder scores higher than 0.4. This includes several emerging and established vaccine candidate antigens such as CTRP, EXP1, TRAP (C-terminal), STARP, GLURP, EBA175 (RIII-V), MSP3, MSP4, MSP6, RALP1, SERA5 and RESA. For these antigens except STARP and RALP1, evidence of balancing selection was consistently observed within disordered regions with only slight variations amongst geographical regions (Fig 12 and S1). Intrinsically disordered regions (IDR) were previously shown to be enriched with predicted linear B-cell epitopes [30]. We found disordered residues within these antigens (i.e. N-term of SERA5 compared to its C-term) contributing to balancing selection (*Tajima’s D* values >1) at most of the analysed geographic area or countries (Fig 12). This suggests IDRs as potential immune targets. However, it has been estimated that IDRs have limited proportion of MHC-binding peptide compared to other domains, which may affect T-cell dependent immune responses [30]. Further *in vitro* and *in vivo* investigations are needed to characterize antigens enriched with disordered proteins as potential vaccine targets.

**Figure 12.**
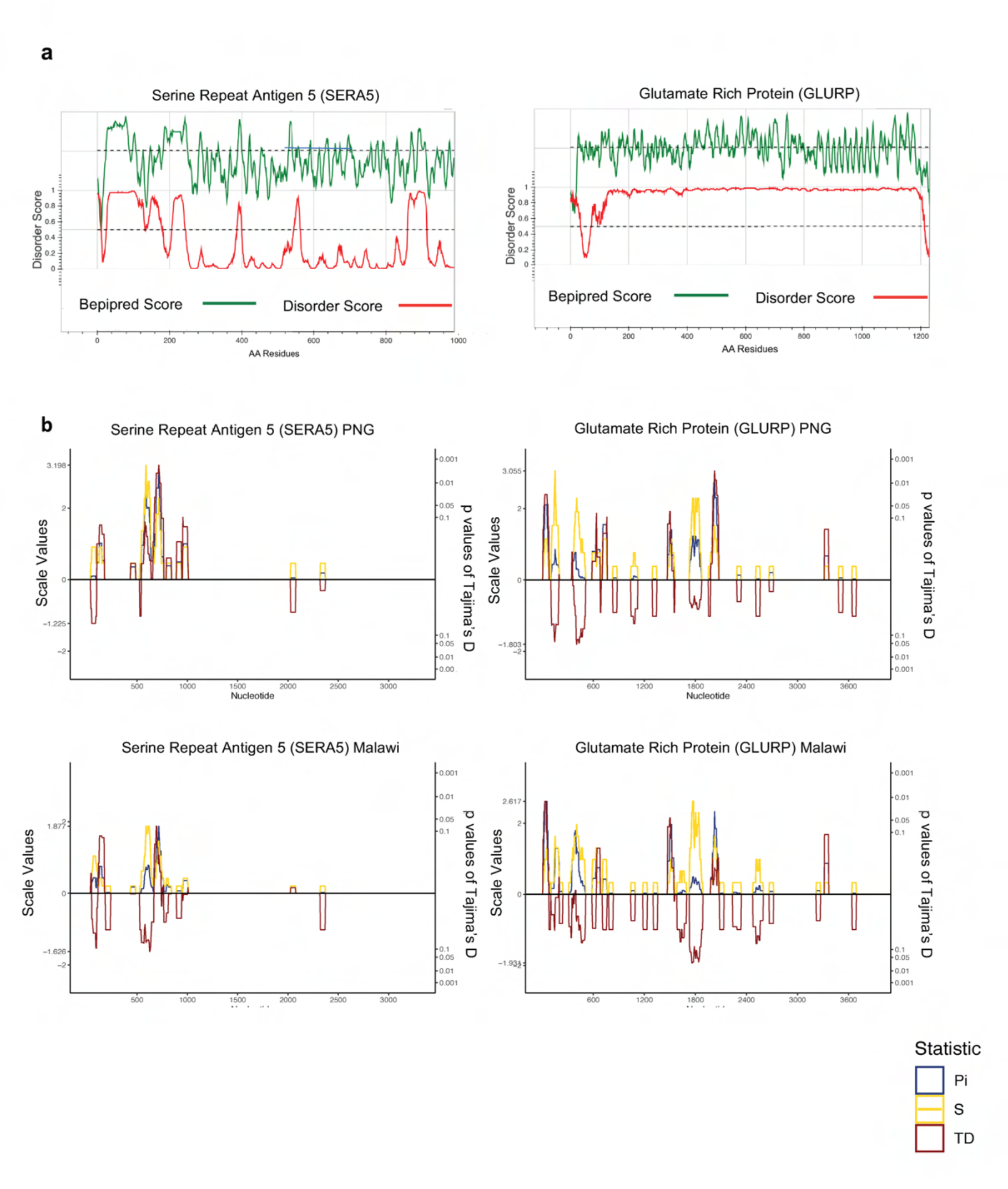
Diversity and selection of SERA5 and GLURP in Asia-Pacific and African regions. a) Computational predictions of protein disorder and B-cell epitopes in SERA5 and GLURP. The green line represents the linear B-cell epitope mapping scores and the red line shows the protein disorder score, respectively. b) Diversity statistics along the sera5 and glurp genes in samples from in Asian-Pacific and African regions, represented by Tajima’s D (red line), nucleotide diversity (blue line) and number of segregating sites (yellow line). It is calculated in the context of linear sequence level based on coding region with the sliding window approach (a window size of 50 bp and a step size of 5 bp). Nucleotide positions based on coding region are shown in the x-axis. Sample sizes: Malawi (n = 106), Ghana (n = 208), Cambodia (n = 405), and PNG (n = 108).

### Antigens with multiple balancing selection hotspots have large numbers of low frequency haplotypes

Analysis of the frequency and relationships amongst amino acid haplotypes revealed many rare haplotypes within African populations, and relatively common haplotypes were detected mostly within Asia-Pacific countries, most likely owing to greater recombination amongst distinct clones in Africa. For antigens with multiple balancing selection hotspots such as CelTOS, TRAP, AMA1, EBA175, MSP1 and GLURP, high haplotype diversity is ubiquitous amongst different countries (Hd > 0.97), and no predominant haplotypes were present in any country. All except CelTOS are non-gametocyte antigens suggesting these antigens as dominant targets of natural host immunity amongst the antigens studied.

Haplotypes were often found in many countries in the dataset (i.e. multiple colours for each node) due to minimal geographic differentiation of each surface antigen. However, clustering of haplotypes based on geography was observed for CTRP, TRAP (ectodomain), CSP (C-term), GLURP, EBA175 (RII), CelTOS, and Pfs48/45. Haplotypes from the same geographical region (Africa or Asian) were more closely connected than those from the other region (Fig 13). Some of these antigens are non-merozoite antigens, consistent with previous observations [7] and may reflect unique adaptations within different geographic areas, or population structure. The haplotype network results were aligned with spatial *D\** analysis for EBA175 (RII), CSP (C-term), CelTOS, and Pfs48/45, finding different selection patterns amongst different populations. Overall, the prevalence of the 3D7 haplotype is low for most of the antigen (Fig 13). Of all analysed antigens, the 3D7 haplotype was prevalent only within CyRPA, MSP1-19, MSP3, MSP4, Pfs48/45, RALP1, RH5, RIPR, STARP, TRAMP, and TREP antigens (Fig 13, Table S1). No 3D7 haplotypes were observed in the dataset for full length EBA175, MSP1 and SERA8 (table S1).

**Figure 13.**
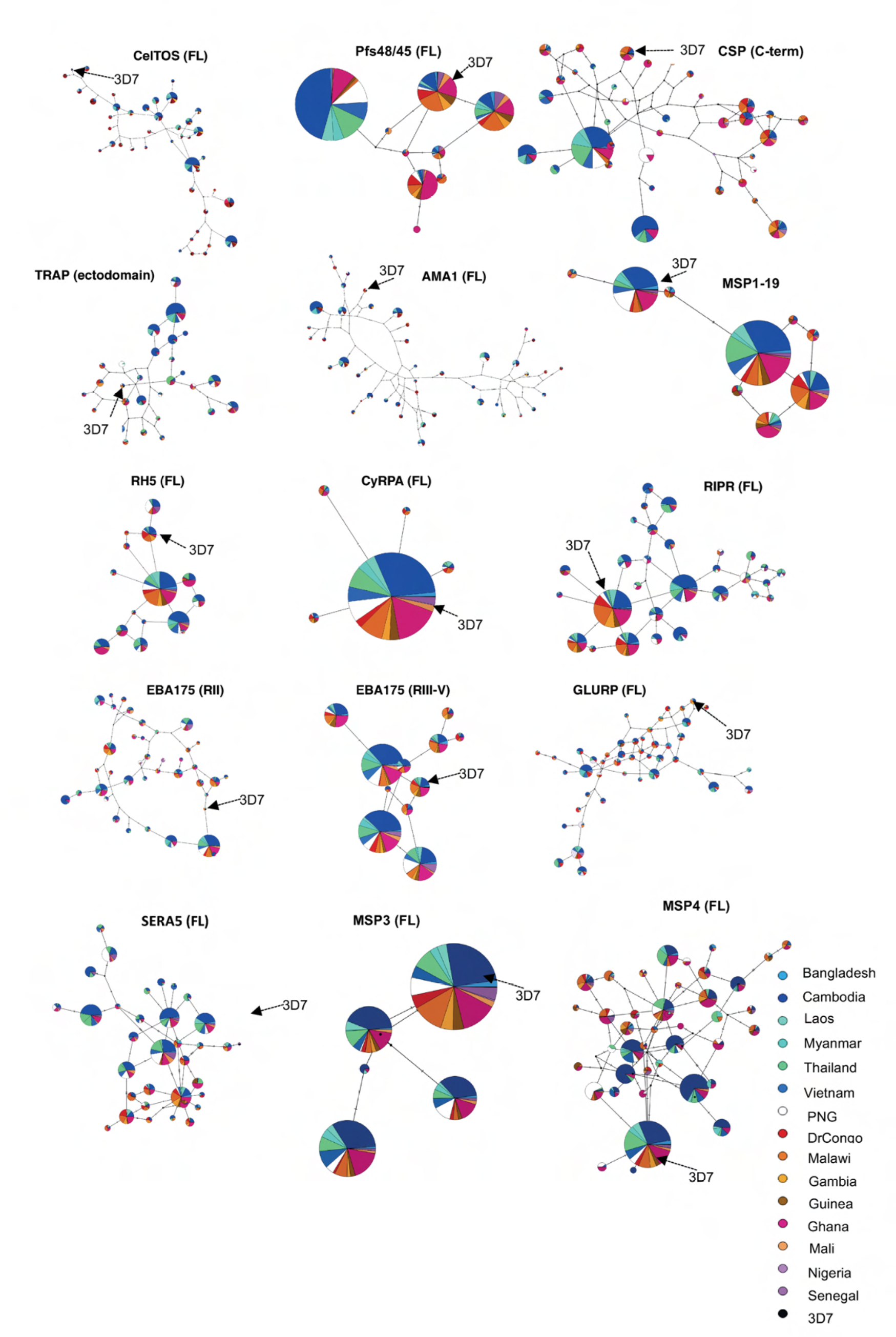
Haplotype network for malaria vaccine antigens. Templeton, Crandall, and Sing (TCS) network summarizing the global diversity of selected antigens using only common haplotypes (> 0.5% of all haplotypes) based on non-synonymous SNPs for full length respective domain of each antigen. Circles represent unique haplotypes, and circles are scaled according to the prevalence of the observed haplotypes. The number of non-synonymous SNP differences between each haplotype was shown by the number of hatch marks on the branches. Vaccine strain 3D7 (arrowed) is included for reference. Results were shown for gametocyte (CelTOS), pre-erythrocytic (CSP (C-terminal), TRAP (Ectodomain), GLURP), and erythrocytic (MSP1_19_, RH5, CyRPA, RIPR, EBA175 (RII), EBA175 (RIII-V), SERA5, MSP3, MSP4) antigens.

## Discussion

Here we have conducted a systematic investigation of the genetic diversity of 23 malaria vaccine antigens using a global dataset of more than 2600 publicly available whole genome sequences covering 15 malaria-endemic countries [19]. Our results demonstrate low frequencies of variants included in vaccines under development for most candidates, and variable diversity and selection amongst the antigens and geographic regions. Overall, the results demonstrate that the current ‘one size fits all’ approach to vaccine development may result in limited vaccine efficacy and variable efficacy across geographic regions [95].

The variable success of malaria vaccines may be due to the high diversity of vaccine candidate antigens [7, 15], and strain-specific immune responses. Merozoite antigens, particularly AMA1, and EBA175 have high haplotype and nucleotide diversity across different populations consistent with strong immune selection. Variable levels of nucleotide diversity for different populations were observed for CSP (C-term) and CelTOS which may result from different transmission intensities. Low nucleotide diversity and moderate haplotype diversity across different populations for STARP, RH5, and RALP1 suggesting that although there is minimal flexibility for accumulation of polymorphisms, immune selection may operate on these antigens to maximise the chance of new infections carrying novel haplotypes. Only limited haplotype and nucleotide diversity was observed within CyRPA, TRAMP, and Pfs48/45 suggesting strong conservation and limited immune selection. For STARP, RH5, RALP1, and CyRPA antigens, low expression throughout the erythrocytic stage (based on abundance of *in vivo* mRNA transcript) suggests minimal opportunities for host evolutionary pressure to be exerted upon those antigens [70].

Previous studies have indicated that parasite antigens are differentially immunogenic [80] and this manifests in varying intensity of immune (balancing) selection. We have detected extremely high balancing selection across different populations for AMA1 and EBA175 (RII), moderate to high balancing selection was found for TRAP (ectodomain) and RIPR. Whereas CyRPA, and SERA5 (C-term) appear to be consistently low diversity, and neutrally evolving in most populations, which implies they have highly conserved epitopes or weak immunogenicity. Variable selection amongst different populations was found for RH5, CSP (C-term), CelTOS, MSP-19 and Pfs48/45 (Fig S9). High-affinity, receptor-specific binding between parasite ligand and host receptor (such as EBA175 – Glycophorin A) or low-affinity general cell binding (such as TRAP – heparan sulphate proteoglycans (HSPGs)) may explain different balancing selection intensity on these antigens amongst populations [81]. The high relative solvent accessibility (RSA) of polymorphic residues on some of these antigens suggests they may be antibody targets, supported by considerable amino acid variability (Shannon Entropy) at host-parasite interfaces, particularly for TRAP (ectodomain), CSP (C-term), AMA1, EBA175 (RII), RH5, Pfs48/45 (6C domain), and CelTOS. All of these antigens have selection hotspots at these interfaces in at least one population.

Hotspots of balancing selection were common within functionally important 3D interfaces of AMA1 and TRAP across populations suggesting sharing of dominant epitopes which may allow the development of broadly efficacious vaccines using a few common variants. On the other hand, we identified differential patterns of selection pressure between African and Asia-Pacific countries at the ligand binding sites of EBA175 (RII), RH5, MSP1-19 and CSP (C-term). Even more discordant patterns between geographic regions were found for gametocyte antigens such as CelTOS and Pfs48/45. This might reflect parasite adaptation to different host environments. For blood stage antigens, EBA175 and RH5, this could be driven by different isoforms of the respective host receptors such as glycophorin-A (GPA) [77] and Basigin. GPA encoded by *GYPA* gene is under selective pressure in different malaria endemic areas, with the MN blood group in particular showing protective associations against malaria in PNG [77, 104]. Associations between different isoforms of BSG with malaria outcomes have not yet been identified however *in vitro* studies have shown a reduced RH5 interaction with isoform 2 of BSG [79]. Despite the very low diversity of RH5 there are multiple common haplotypes, with strong balancing selection at the BSG binding site. However, this balancing selection was only found within Asia-Pacific populations, and not in Africa. This suggests that parasites may have adapted RH5 to an as yet unidentified BSG isoform in the region, or that some other factor might be contributing to selection at this BSG binding interface. Similarly, CSP (C-term) and CelTOS shows unique selection hotpots on the 3D structure as these proteins are predominantly expressed in the sporozoites, this might reflect adaptation to different Anophelene species in Africa and Asia. This geographically variable selection was not easily observed with sliding window analysis of linear sequences showing the enhanced resolution of the 3D analysis.

Immune responses may also be generated to conserved regions of antigens [82] such as that in the novel universal influenza B-cell vaccine approach [83]. This would increase vaccine efficacy for diverse strains by eliciting broadly cross-reactive functional immune responses such as neutralising antibodies directed at highly conserved regions. Conserved regions of malaria vaccine candidate antigens have been revealed within RH5, CyRPA, or SERA5 (C-terminal) (Fig S8) and a lack of selection pressure in the C-terminal region of SERA5 and CyRPA. Therefore, not only does this study catalogue and characterise global malaria vaccine antigen diversity, it can also guide the identification of conserved regions. This study therefore provides critical practical information for the next generation of malaria vaccines.

Limited investigations have been performed to understand the immunogenic properties of disordered proteins. Lower immunogenicity of a disordered region compared to a structured region within MSP2 has been observed [85] suggesting that disordered proteins fail to activate the production of high affinity, specific antibodies, because they adopt diverse conformations depending on their ligands [86]. Therefore, polymorphic hotspots on disordered proteins may act as a ‘smoke screen’ by diverting protective immune responses from more ordered targets. Nonetheless, numerous B-cell epitopes have been found within disordered regions [30] and many of these appear to elicit functional immune responses [87–91]. For instance, the protective effect of RTS,S appears to be predominantly mediated by generation of antibody responses to the disordered central NANP repeat region [91]. Our observation of immune-mediated selection pressure in the N-terminal disordered region of SERA5, is also supported by *the finding that in vitro* parasite growth was inhibited by anti-SERA5 antibodies raised in squirrel monkeys (*Saimiri sciureus*) [90]. In addition, we found balancing selection in the disordered EBA175 RIII-V domain which may facilitate the molecular binding of GYPA to EBA175 RII [92, 93]. Future work could extend to characterise disordered proteins into different subclasses based on their immunogenicity.

This study presents a detailed analysis of the diversity and immune selection of 23 candidate vaccine antigens and a framework for analysis of further antigens which has already contributed critical information about the diversity of novel vaccine candidates [96, 98]. In addition, the incorporation of sequence-based metrics such as Tajima’s D into protein structures enhances biological understanding, as evidenced by the finding of unique patterns of immune selection in different geographic areas. *In silico* findings from our study support *in vitro* and *in vivo* studies such as the finding of monoclonal antibody binding targets and might uncover novel targets of host immunity. Thus, the implementation of presented data analysis and the development of population genetic tools was crucial for our analyses. The approach used in this work is also applicable to a wide variety of problems from different organisms.

## Materials and Methods

### Data Sources

WGS of *P. falciparum* clinical isolates (n = 2512) from multiple locations of Asia and Africa including Bangladesh, Cambodia, Laos, Myanmar, Thailand, Vietnam, Congo (DRC), Malawi, Gambia, Ghana, Guinea, Mali, Nigeria and Senegal. were obtained from the MalariaGEN Pf3k project release 5.1 as unmapped BAM (uBAM) files [18]. WGS of *P. falciparum* isolates from PNG (n = 156) were also obtained. All samples were sequenced using Illumina short read technology in the Sanger Research Institute, UK, and the Broad Institute, Boston, USA using different library preparations including different DNA extractions and enrichment methods [18, 20]. The sequences from these 15 countries cover four broad geographical regions (Southeast Asian, Central Africa, West Africa, and Oceania) with different malaria transmission intensity.

### Processing and Analysis of Whole Genome Sequence data

*P. falciparum* read alignments from 2668 samples were mapped to an indexed 3D7 reference genome using uBAM files as input. We followed the standard best practice from *Genome Analysis Toolkit (GATK)* version 4.0.12.0 implemented in *nextflow* (https://github.com/gatk-workflows/gatk4-germline-snps-indels) with some minor changes as follows: (i) running the pipeline twice - the first without base quality score recalibration (BQSR) and (ii) using the pass variants from the first run for BQSR and hard filtering of variants instead of variant quality score recalibration (VQSR) as there is no external variant callset available. Variants were determined using GATK’s *haplotypeCaller* to generate haploid genotype calls (*P falciparum* blood stages are haploid) and all isolates were joint genotyped using gvcf files. Variants that had been processed with the GATK pipeline were functionally annotated with *SnpEff for* genomic variant annotations and functional effect prediction (version 4.3T) [21]. We restricted our analyses to single nucleotide polymorphisms (SNPs) from the 14 nuclear chromosomes only, excluding indels and variants from hypervariable regions, as they evolve by different mechanisms to SNPs and have much less impact on antigen genes (Fig S1). To obtain a high-quality variant dataset for downstream analyses, we further developed stringent variant filters using GATK’s SelectVariants and VariantFiltration modules such as read coverage statistics, relative distribution of reads [22] and multiplicity of infection (MOI) (Fig S1). Variants were removed if they met at least one of the following filtering criteria: QD < 20.0, MQ < 50.0, MQRankSum < -2 (if annotated), ReadPosRankSum (less than -4 or greater than 4 if annotated), SOR > 1. Further filtration was performed based on read depth, and missingness (Fig S1). We estimated the clonal proportions for each sample using R package, *moimix* (available at https://github.com/bahlolab/moimix), a robust alternative (B)-allele frequency-based estimation. We only kept data from samples with single infections (MOI= 1) or two infections (MOI = 2; i.e. Fws > 0.80) [4]. MOI = 2 isolates were treated as diploid genomes permitting heterozygous genotype calls (Fig S2). Multi-allelic sites were also permitted in our bioinformatics framework (Fig S1). All samples that had F_ws_ greater than 0.90 were treated as MOI=1. For MOI=2 samples, phasing of haplotype was performed using read-depth and B-allele frequency (Fig S1). Thus, the final dataset contained high quality data from 948 MOI=1 and 551 MOI=2 isolates. Nucleotides that did not have clear major or minor alleles were classified as ‘undefined’ bases (Fig S1). Variants within gene coding regions were extracted in FastA format using a custom R-script (https://github.com/myonaung/Pf3K_Scripts). We found a large proportion of field isolates from African regions (greater than 30%) with high MOI (>2), especially in Malawi and Ghana, likely reflecting high transmission intensity in those areas. However, the sample size in the final dataset (n = 1499) remained high, even after removing these samples. After processing the sequence data, identifying high quality variants and removing complex infections, gene sequences were extracted and compiled. The diversity and evolutionary signatures found within each gene was analysed. In addition, we mapped polymorphisms and selection signatures onto available 3D protein structures to investigate the spatial distribution of polymorphic residues and possible functional consequences.

### Analyses

Antigens were selected based on their inclusion as vaccine candidates in the WHO Malaria Rainbow Table [23] or whether they had been previously identified as novel candidates based on previous literature [12]. Details of regions analysed, and other details are summarised in Table 1. The raw fastA output for each gene was further edited using customized python script (https://github.com/myonaung/Pf3K_Scripts) by removing sequences that contain undefined or low-quality bases and sequences arising from minor clones in the co-existence of different genotypes (phased by read depth (Fig S1)). FastA formatted files of respective genes can be found in https://github.com/myonaung/Pf3K_Scripts/tree/master/Raw_FASTA. As Illumina sequencing is subject to some miscalled bases, singleton variants (i.e. minor alleles seen only once in the entire dataset) were assumed to be artefacts and converted back to reference alleles to prevent false positive variants. The presence of at least two isolates with the same minor allele was considered independent validation of the existence of these alleles in natural parasite populations. Apart from removing singletons, no other minor allele frequency threshold was applied to the dataset. In the case of multiple clones being identified (0.90 ≥ Fws > 0.80), only the defined major alleles were included for further analyses. Samples collected from the same country were combined and assumed to have been sampled from a single population. Population genetic parameters were calculated using a customized in-house R package (source code available at https://github.com/BarryLab01/vaxpack). This program takes aligned multi-fastA files excluding intronic regions as input, alongside reference sequence of the same length. As measure of diversity we defined the polymorphisms for each antigen by the number of polymorphic sites (S), the number of synonymous (dS) SNPs, number of nonsynonymous (dN) SNPs, dN-based haplotypes, the Nei’s nucleotide diversity (π) calculated as

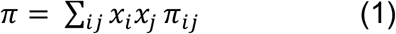

where *x_i_* and *x_j_* are the respective frequencies of the i^th^ and j^th^ sequences, π_*ij*_ is the number of nucleotide differences per nucleotide site between i^th^ and j^th^ sequences [24]. The haplotype diversity (Hd) was defined as

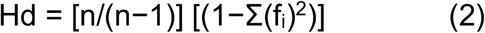

where n is the sample size and f is the frequency of the i^th^ allele. Normalized Shannon Entropy for each amino acid residue from the protein sequence was calculated using

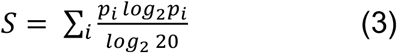

where S is the normalized Shannon Entropy, and p_i_ is the frequency of i^th^ amino from the specific position [24]. The normalized Shannon Entropy is a site-specific diversity measure ranging between 0 to 1 where 0 represents the conservation of a specific amino acid position, and 1 represents an even distribution of all possible standard 20 amino acids (random distribution) at the specific position. Immune mediated selection due to parasite adaptations to evade antibody recognition of dominant epitopes on a particular antigen is indicated by the presence of balancing selection. To measure selection, *Tajima’s D* (*D*) statistics were calculated [24, 25]. Under neutral conditions, *Tajima’s D* values are expected to be approximately zero [26, 27], positive *D* values indicate an excess of intermediate frequency polymorphisms most likely due to balancing (immune) selection, and negative values indicate purifying (directional) selection. However, measurement of *Tajima’s D* across genetically differentiated populations with varying allele frequencies (i.e. due to population structure) can produce false positive signals of balancing selection. Thus, all *Tajima’s D* analyses were calculated separately for each country. Analyses were first carried out with the complete global dataset (n = 1499 samples) for all antigens from Table 1 to investigate the global range of diversity of each antigen as well as the frequency of reference (3D7) haplotypes (frequently used in vaccine development), while the population specific dataset was used to investigate the range and distribution of diversity for countries (23 antigens). RON2 (PF3D7_1452000) was excluded from analyses as no polymorphism was found. The full-length antigen genes as well as functionally important domains were included in the country level.

For each analysed antigen gene, we investigated relationships amongst haplotypes by constructing haplotype networks using the Templeton, Crandall, and Sing (TCS) method in *PopArt* version 1.7 [28]. To focus the analysis on antigen diversity, we based our analysis on non-synonymous polymorphism (via amino acid translation) as synonymous polymorphisms do not change the protein structure. It is very important to note that haplotypes are simply the combination of polymorphic sites without a particular weight upon a specific or functionally important polymorphic residue. All nonsynonymous SNP haplotypes with a frequency greater than 0.5% of total population were included in the analysis using ‘Nexus’ file format as input. The TCS network was constructed using an agglomerative approach. This allows for visualization of the relationships amongst haplotypes and their geographic distribution. Generally, less frequent recombinant haplotypes connect major (high frequency) haplotypes.

The Relative Solvent Accessibility (RSA) derived from the solvent accessible surface area (ASA) was predicted for all antigens for each amino acid residue based on primary 3D7 protein sequence using neural network based NetSurfP1.1 program [29, 102]. It is given by the following equation [29, 102]:

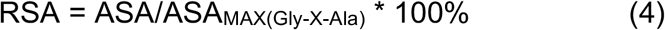

RSA values for structured regions with known PDB or homology-modelled structures were calculated with DSSP [103] using maximum allowed solvent accessibilities of residues from Tien *et al.* 2013 [102]. Therefore, RSA represents the ratio of parts of a biomolecule exposed to solvent or accessible surface area (ASA) of a given residue observed in the three-dimensional state over maximum surface area of a residue with potential exposure to solvent (ASA_MAX_) within an extended tripeptide flanked with either glycine or alanine residue. The data was stored in SQLite database using RSQlite package on RStudio.

Selection pressure may apply to residues that are non-continuous on the linear sequence but proximal in 3D space. Therefore, application of a more accurate spatially derived approach to compute *Tajima’s D\** and nucleotide diversity is very applicable. We chose the radius of 15 Å to reflect the ideal potential antibody antigen interaction as used by Guy *et al.* (2018) [8, 9]. However, our spatial averaging approach was limited to the availability of 3D coordinates for each amino acid residue, and therefore were not applied to highly disordered regions or proteins.

### Protein Structures

For each antigen included in Table 1, the 3D structure for full length or specific domains was first derived from the Protein Data Bank (PDB) from the Research Collaboratory for Structural Bioinformatics (RCSB) website (www.rcsb.org). For each antigen that does not have experimentally determined or complete structure, the possibility of comparative structure modelling is determined based on the distribution of intrinsic disordered regions available at https://plasmosip.burnet.edu.au/submission [30]. Except for AMA1, which was modelled manually, template-based models of 3D7 reference strain for these antigens were predicted using *ModPipe*, an automated software pipeline that utilises the program *MODELLER* [31] for the generation of comparative proteins based on previous studies [9]. We used a manually modelled structure of AMA1, which includes full domains I-III based on a combination of *P. falciparum* and *P. vivax* templates according to previously published work [32]. The modelled structures are assumed to be reliable if the *ModPipe* Quality Score (MPQS) is greater than 1.1 This includes for antigens – EBA175 (RII), MSP1_19_, and CelTOS which is 45% sequence identity to *P. vivax* CelTOS structure (PDB Code: 5TSZ)[33]. These structure models are accessible via *ModBase* (https://modbase.compbio.ucsf.edu/). The *BiostructMap* Python package available at https://github.com/andrewguy/biostructmap was used to calculate nucleotide diversity (π), and *Tajima’s D* with consideration of spatial information using a 3D sliding window with a radius of 15 Å (*D\** = spatially derived D). Since the spatial *Tajima’s D* (*D*\*) calculation also considers the spatial information of each amino acid residue from the protein, the results represent accurate detection of selection pressures arising at the level of protein structure but might be variant from the traditional linear *D* calculation as shown in Guy *et al.* (2018) [8, 9]. *D\** scores of 1.0 to 1.9 are considered as moderately high and *D\** scores of above 2 are high. Protein structures and features were visualised using *PyMol* with feature values saved into the B-factor column of a PDB file and using the *spectrum* command. We estimated disordered scores using the PlasmoSIP online tool (https://plasmosip.burnet.edu.au), which contains precalculated results from DISOPRED3 [62]. Due to the lack of well-defined three-dimensional structure for antigens with extensive disordered regions, we calculated linear *Tajima’s D* values to determine balancing selection.

## Data Availability

All data used in this study are publicly available

## Acknowledgements

We are grateful to the communities and researchers of malaria endemic countries that enabled the collection and available of the *P. falciparum* genome sequences used in this project, made publically available through the MalariaGEN *P. falciparum* Community Project. We also thank Z. Pava for critical reading of the manuscript. The project was funded by the Australian National Health and Medical Research Council (NHMRC) Project Grant 1161066. MB and IM were supported by NHMRC Research Fellowships (GNT1102971, GNT1155075). This work was made possible through Victorian State Government Operational Infrastructure Support and Australian Government NHMRC IRIISS.

## Supporting Materials

**Figure S1.**
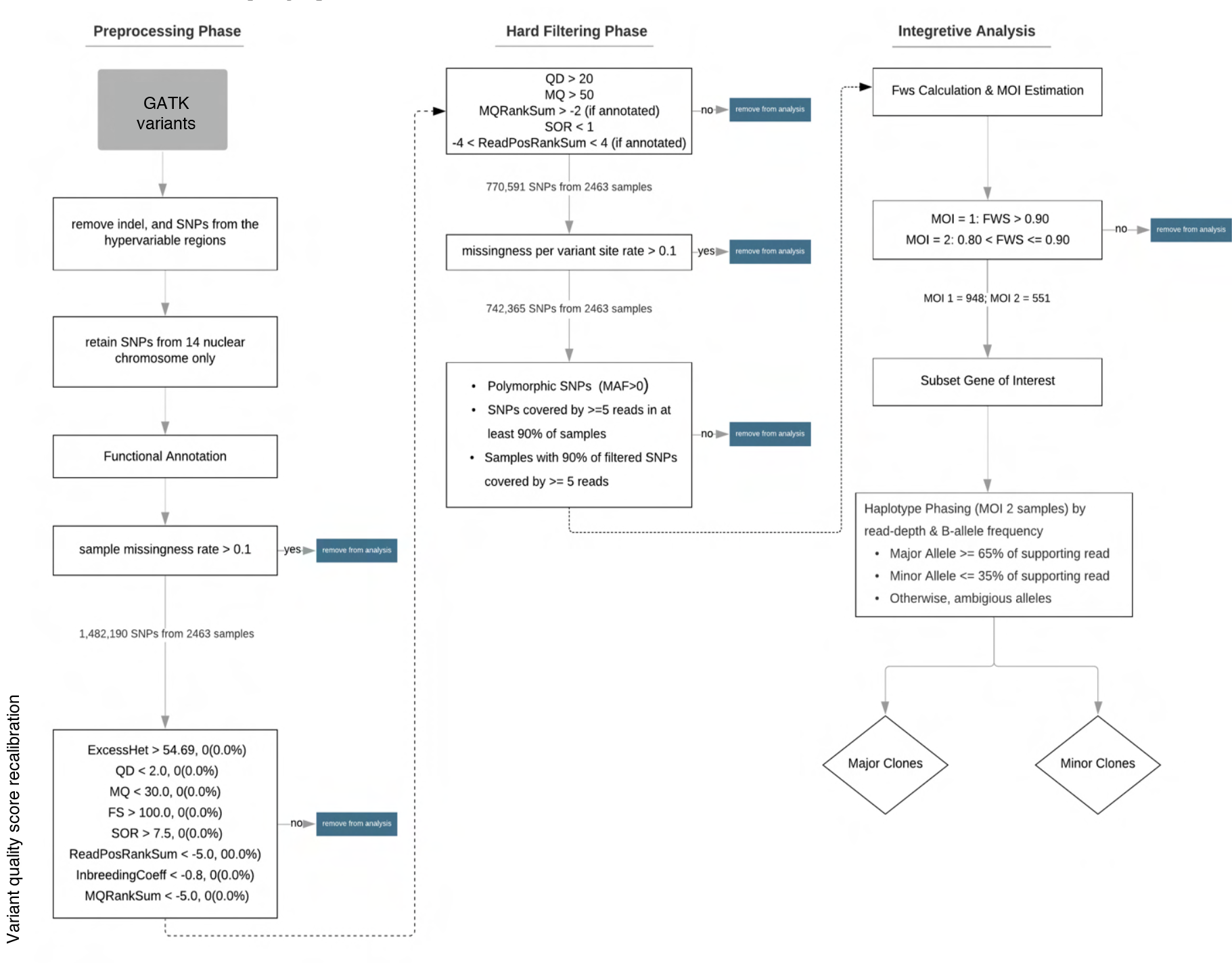
Variant Processing Framework. A pre-processing phase specifies GATK’s Haplotype caller’s SNPs variants from the regions of interest and performs quality control on these variants. The hard-filtering phase selects only high-quality variants from the variants that passed the pre-processing phase. The integrative phase removes polyclonal (> MOI 2), performs haplotype phasing, and extracts sequences for gene of interest.

**Figure S2.**
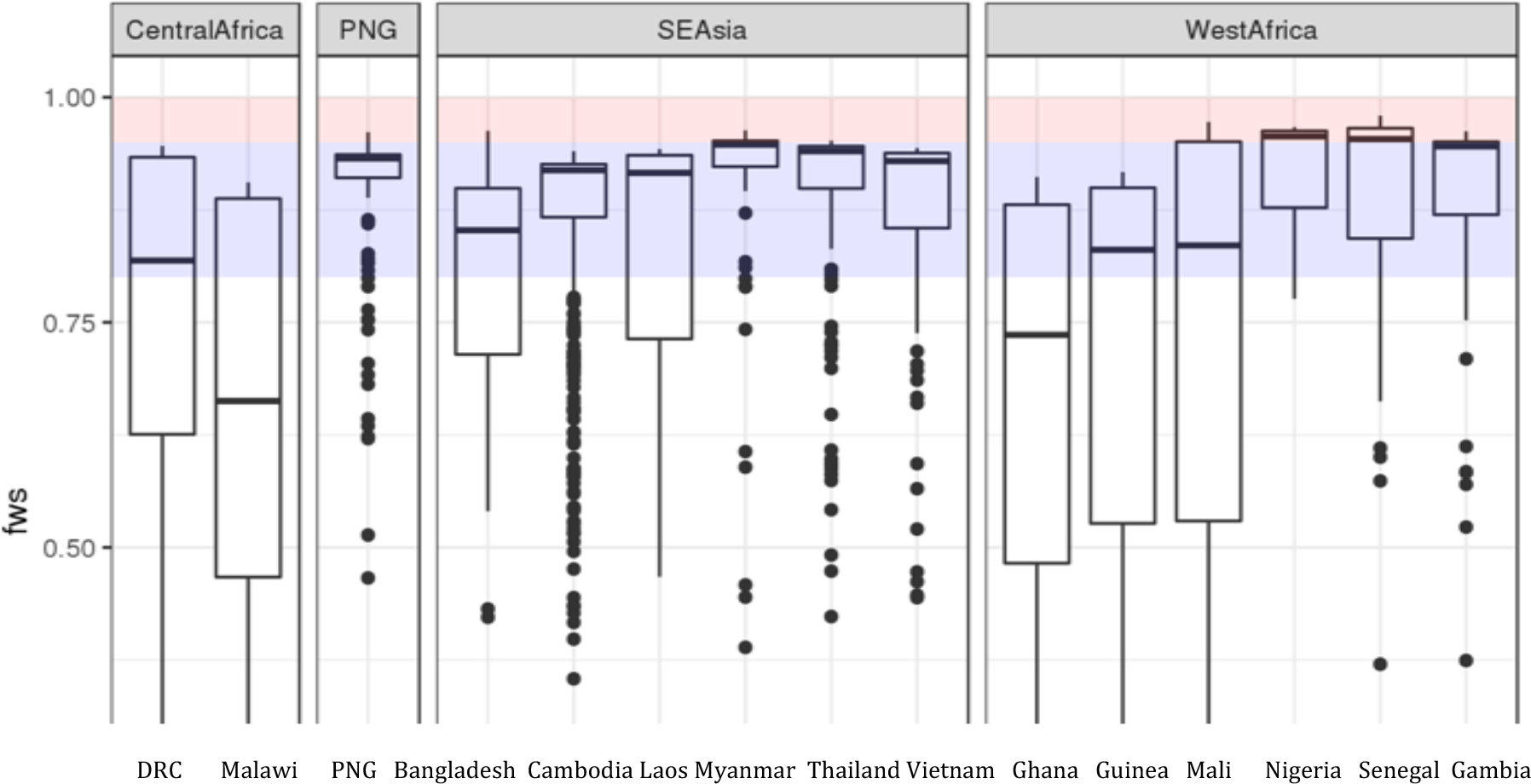
F_ws_ output from moimix R-package for each country. F_ws_ > 0.90 assumed as MOI 1 isolates are highlighted in red, and 0.90 ≥ Fws > 0.80 assumed as MOI 2 isolates are highlighted in blue. Samples with F_ws_ below 0.80 are excluded from the analysis.

**Table S1:** Antigen Diversity Summary (Full length or specific domain)

| Antigen Genes | Length<br>(base pair) | Global NonSyn/Syn<br>SNP Ratio (NS/SP) | Global<br>Number of AA<br>Haplotype | Global haplotype<br>Diversity (Hd)<br>(Median) | Global nucleotide<br>Diversity (Median) ($\pi$ *<br>10<sup>-3</sup>) | Global 3D7<br>Proportion (%) | Sample Size (n)* |
| --- | --- | --- | --- | --- | --- | --- | --- |
| *Csp* | 1194 | 3 | 152 | 0.9 | 4.43 | 1.03 | 1100 |
| *Csp (C-terminal)* | 249 | 31 | 110 | 0.91 | 19.32 | 2.97 | 1100 |
| *Ctrp* | 6345 | 2.8 | 212 | 0.78 | 0.66 | 0.07 | 1389 |
| *Trap* | 1725 | 23 | 563 | 0.98 | 7.06 | 0.41 | 1468 |
| *Trap (Ectodomain)* | 598 | 1.69 | 105 | 0.95 | 9.51 | 9.26 | 1468 |
| *Exp1* | 489 | 14 | 25 | 0.73 | 2.21 | 4.58 | 1445 |
| *Starp* | 1785 | 6 | 50 | 0.63 | 0.56 | 56.68 | 1452 |
| *Trep* | 10632 | 3.41 | 484 | 0.96 | 0.50 | 4.8 | 1453 |
| *Glurp* | 3702 | 4.04 | 383 | 0.98 | 2.44 | 0.83 | 1450 |
| *Ama1* | 1869 | 11.78 | 447 | 0.97 | 12.87 | 0.6 | 1478 |
| *Eba175 (FL)* | 4509 | 10.67 | 340 | 0.96 | 2.07 | 0 | 1477 |
| *Eba175 (RII)* | 1848 | 3.88 | 156 | 0.93 | 3.36 | 0.27 | 1477 |
| *Eba175 (RIII-V)* | 1477 | 0.91 | 82 | 0.57 | 1.07 | 34.3 | 1477 |
| *Rh5* | 1581 | 3.67 | 29 | 0.79 | 1.21 | 6.81 | 1499 |
| *Ripr* | 3261 | 5.12 | 108 | 0.91 | 0.78 | 22.56 | 1499 |
| *Cyrpa* | 1089 | 8 | 10 | 0.12 | 0.12 | 94 | 1485 |
| *Msp1* | 5163 | 6.16 | 603 | 0.98 | 4.3 | 0 | 1040 |
| *Msp1-19* | 277 | 3 | 12 | 0.64 | 5.98 | 23 | 1040 |
| *Msp3* | 1065 | 1.33 | 27 | 0.72 | 3.38 | 42.3 | 1202 |
| *Msp4* | 819 | 18.5 | 131 | 0.93 | 3.96 | 20.6 | 1358 |
| *Msp6* | 1116 | 22 | 44 | 0.75 | 2.59 | 39.9 | 1281 |
| *Ralp1* | 2250 | 2.86 | 26 | 0.52 | 0.30 | 55.3 | 1348 |
| *Resa* | 3258 | 3.36 | 145 | 0.93 | 0.95 | 1.13 | 1187 |
| *Sera5* | 2994 | 4.45 | 176 | 0.93 | 2.12 | 1.25 | 1356 |
| *Sera8* | 2040 | 9.5 | 198 | 0.93 | 1.73 | 0 | 1474 |
| *Tramp* | 1059 | 0.67 | 3 | 0 | 0.11 | 99.2 | 1491 |
| *Pfs48/45* | 1347 | 7 | 13 | 0.5 | 0.94 | 16.16 | 1480 |
| *Pfs48/45 (6C)* | 415 | 4 | 7 | 0.5 | 2.07 | 26.4 | 1480 |
| *Celtos* | 549 | 14.67 | 226 | 0.95 | 11.88 | 0.07 | 1488 |

\*Different sample size amongst antigens due to further quality filtrations based on individual gene

**Figure S3.**
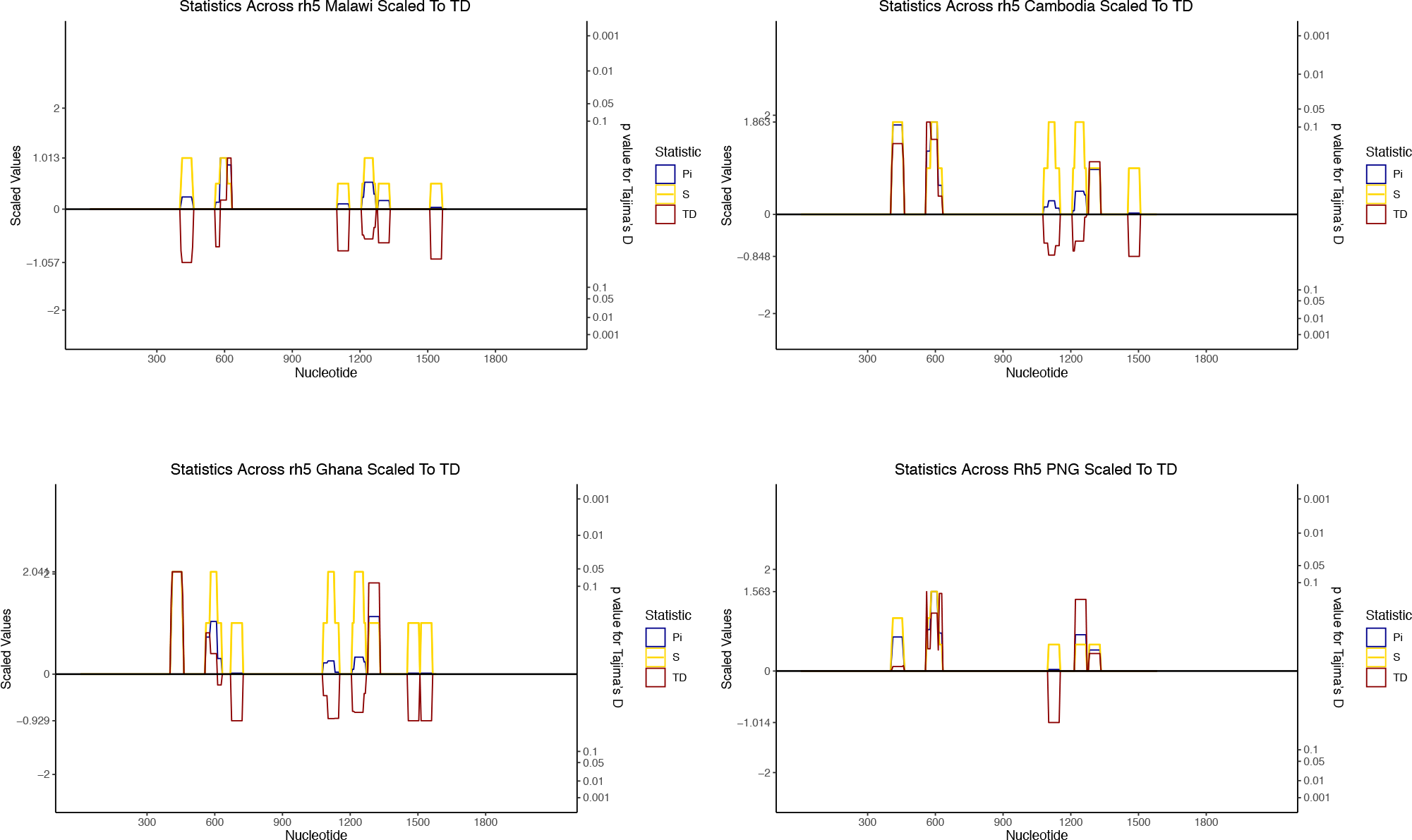
Polymorphism and selection of full-length rh5 in the context linear sequence level for different populations. The sliding window analyses (a window size of 50 bp and a step size of 5 bp) calculated for segregation sites (S, yellow lines), nucleotide diversity (π, blue lines) and Tajima’s D (D, red lines) for each geographic area or country. The results were plotted together and scaled to Tajima’s D values. Nucleotide positions based on coding region are shown in the x-axis. The significant values for Tajima’s D was determined based on sample size.

**Figure S4.** Geographically varied selection pressure for SERA5. Tajima’s D (D*) calculation for geographic area or countries from Asia-Pacific and African regions for SERA5 (C-terminal) with incorporation of protein structural information using 15°A window. The structured region of SERA5 based on experimentally defined structure PDB code: 2WBF was used. The structure was coloured according to D* scores mapped to each residue with undefined D* were shown in grey.Only Malawi (n= 106), and PNG (n=108) populations were shown.

**Figure S5.** Geographically conserved spatially derived nucleotide diversity for full-length AMA1. Nei’s nucleotide diversity calculation for geographic area or countries from Asia-Pacific and African regions for AMA1 with incorporation of protein structural information using 15°A window. Structure was coloured according to nucleotide diversity mapped to each residue. Sample size for each respective population are as follows: Malawi (n= 139), Ghana (n=243), Cambodia (n=433), and PNG (n=112). Similar to selection pressure (determined by D*), silent face of AMA1 has low nucleotide diversity.

**Figure S6.**
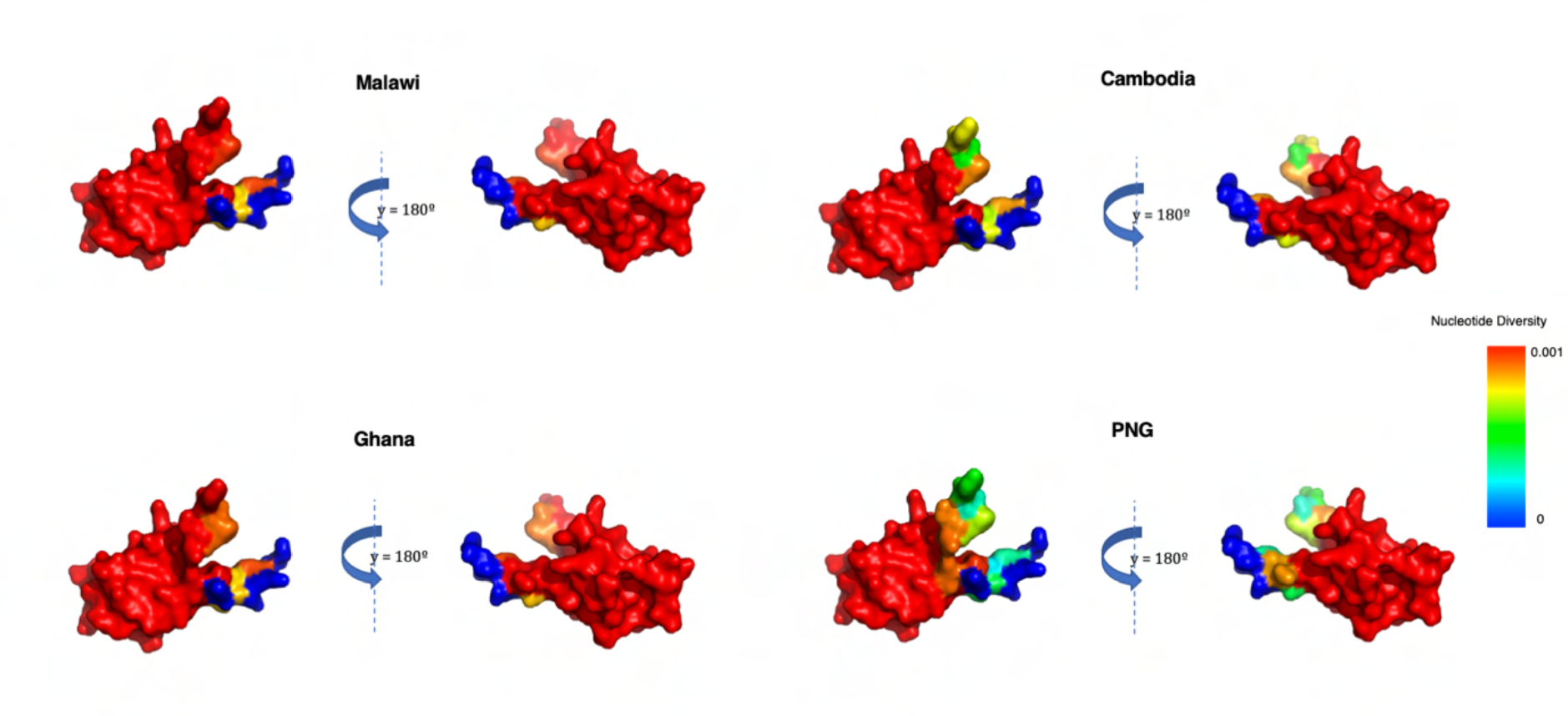
Geographically variable spatially derived nucleotide diversity for CSP (C-term). Nei’s nucleotide diversity calculation for geographic area or countries from Asia-Pacific and African regions for CSP (C-term) with incorporation of protein structural information using 15°A window. Structure was coloured according to nucleotide diversity mapped to each residue. Sample size for each respective population are as follows: Malawi (n= 135), Ghana (n= 223), Cambodia (n=431), and PNG (n=111).

**Figure S7.**
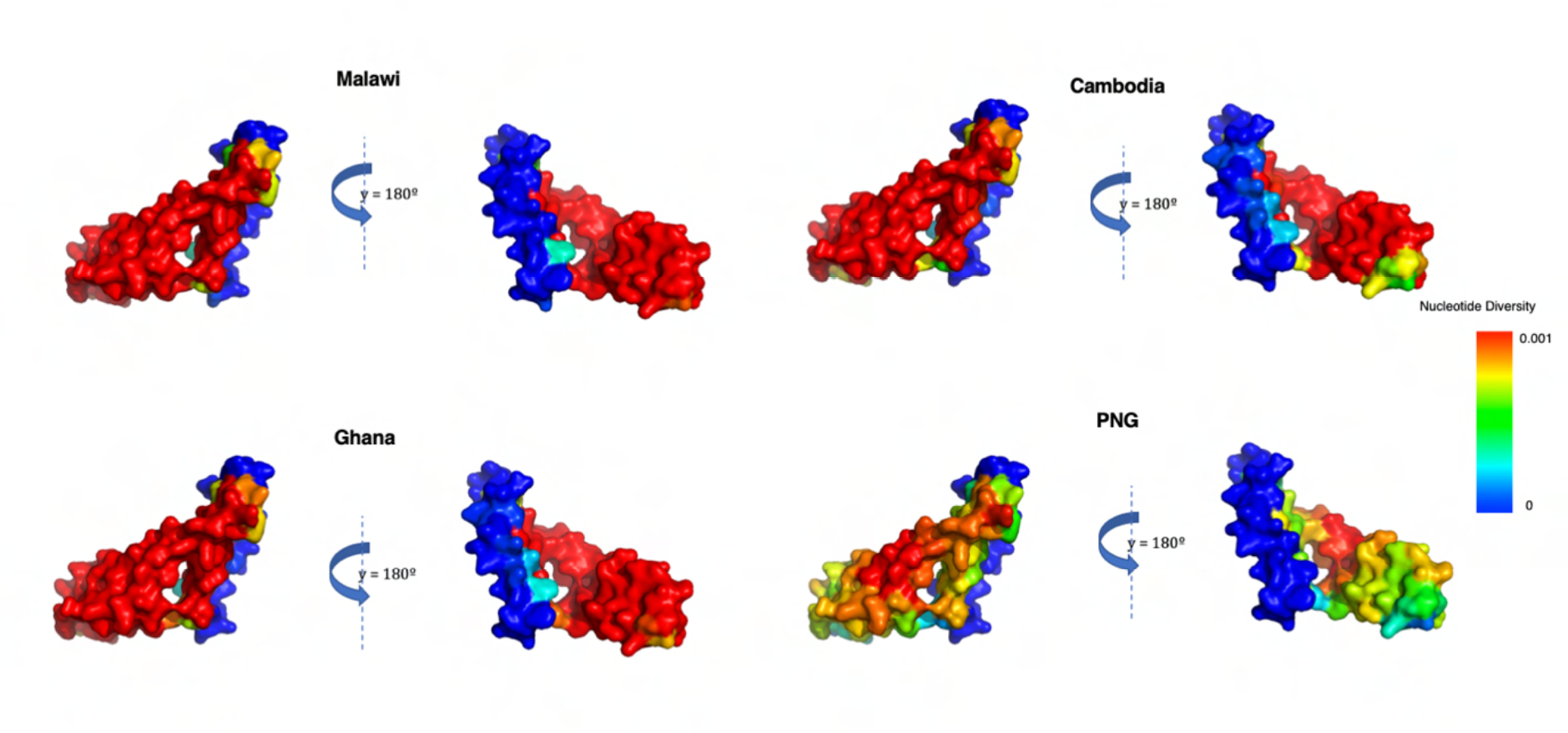
Geographically variable spatially derived nucleotide diversity for CelTOS. Nei’s nucleotide diversity calculation for geographic area or countries from Asia-Pacific and African regions for CelTOS with incorporation of protein structural information using 15°A window. Structure was coloured according to nucleotide diversity mapped to each residue. Sample size for each respective population are as follows: Malawi (n= 142), Ghana (n=245), Cambodia (n=433), and PNG (n=112).

**Figure S8.**
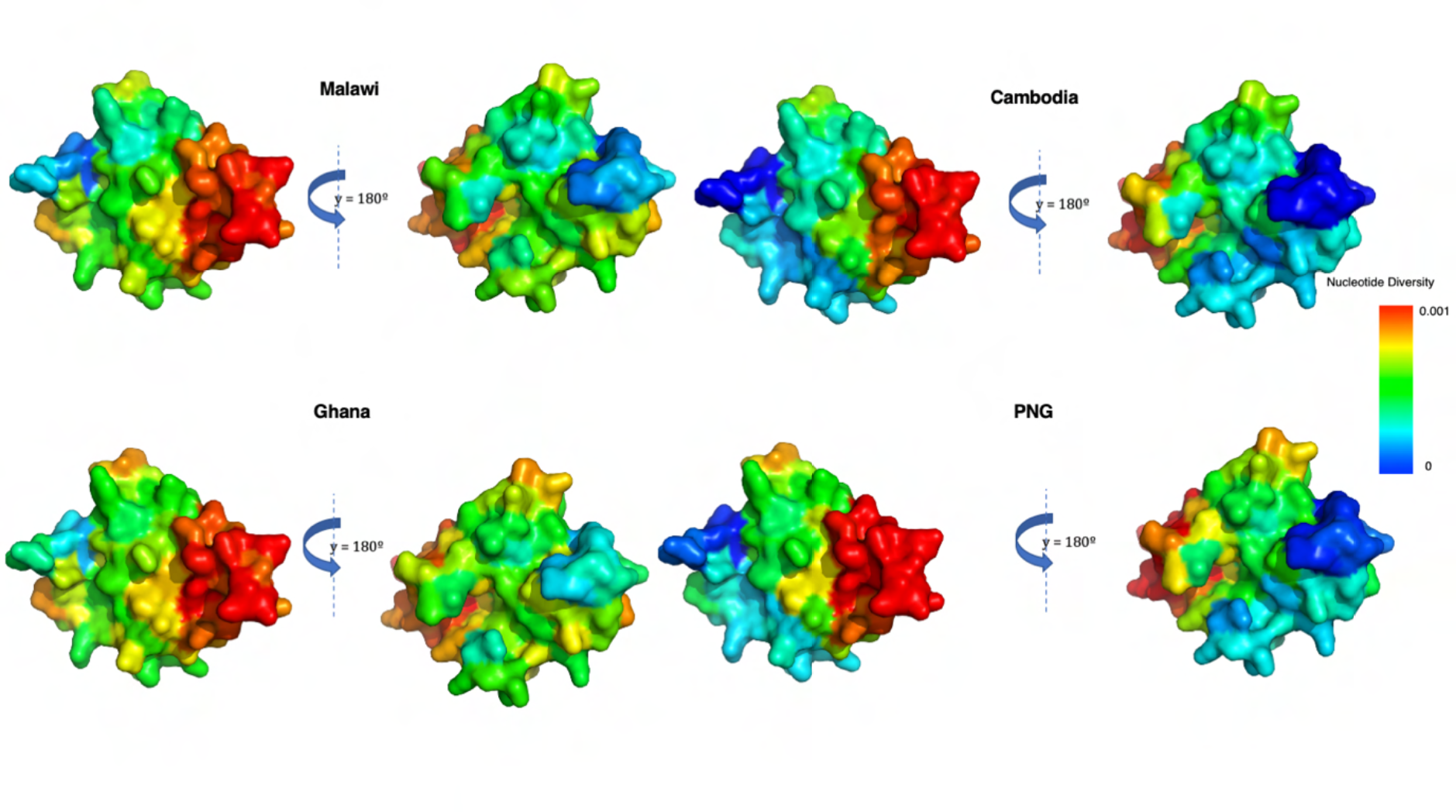
Geographically variable spatially derived nucleotide diversity for MSP1-19. Nei’s nucleotide diversity calculation for geographic area or countries from Asia-Pacific and African regions for MSP1-19 with incorporation of protein structural information using 15°A window. Structure was coloured according to nucleotide diversity mapped to each residue. Sample size for each respective population are as follows: Malawi (n= 101), Ghana (n=183), Cambodia (n=270), and PNG(n=72).

## Geographically variable selection for Pfs48/45

Consistently, moderately *D* scores (1.5 – 2) were observed in the region corresponding to nucleotide residues 780 – 800 in most of the populations (Figure S9). In addition, nucleotide residues 900 – 100 were also under balancing selection within African populations (moderately high *D* of 1.7) (Figure S9). Of these, corresponding amino acid residues N299 and N303 are known to be involved in N-linked glycosylation [3]. Patterns of selection were varied amongst observed populations. However, due to limited number of Pfs48/45 non-synonymous polymorphisms driving these patterns, the results should be interpreted with care.

**Figure S9.**
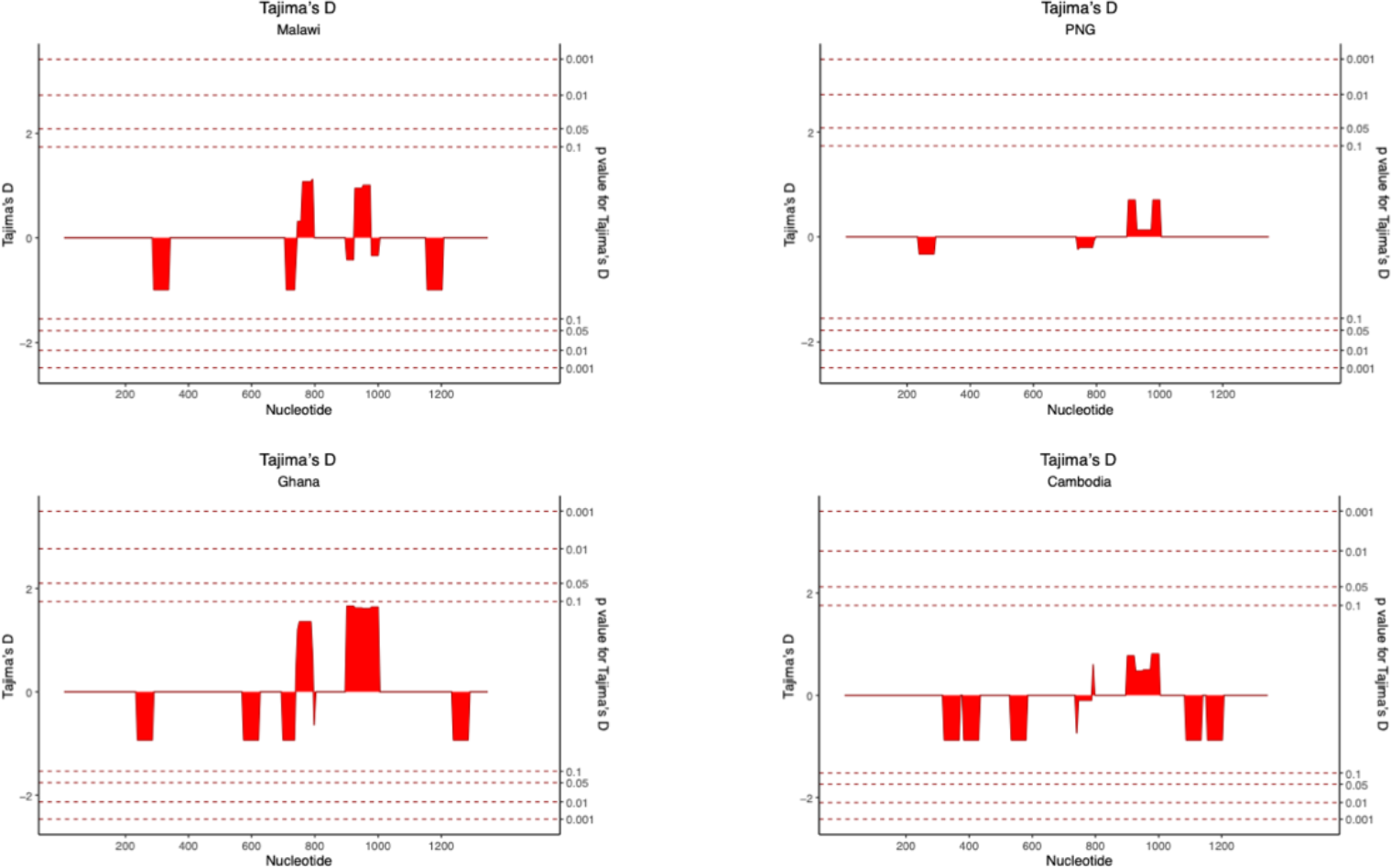
Geographically varied selection pressure for Pfs48/45. The sliding window analyses (a window size of 50 bp and a step size of 5 bp) calculated for Tajima’s D (D, red lines) for each population. Nucleotide positions based on coding region are shown in the x-axis. Significant value for Tajima’s D was determined by sample size. Sample size for each respective population are as follows: Malawi (n= 142), Ghana (n=247), Cambodia (n=433), and PNG (n=112).

**Figure S10.**
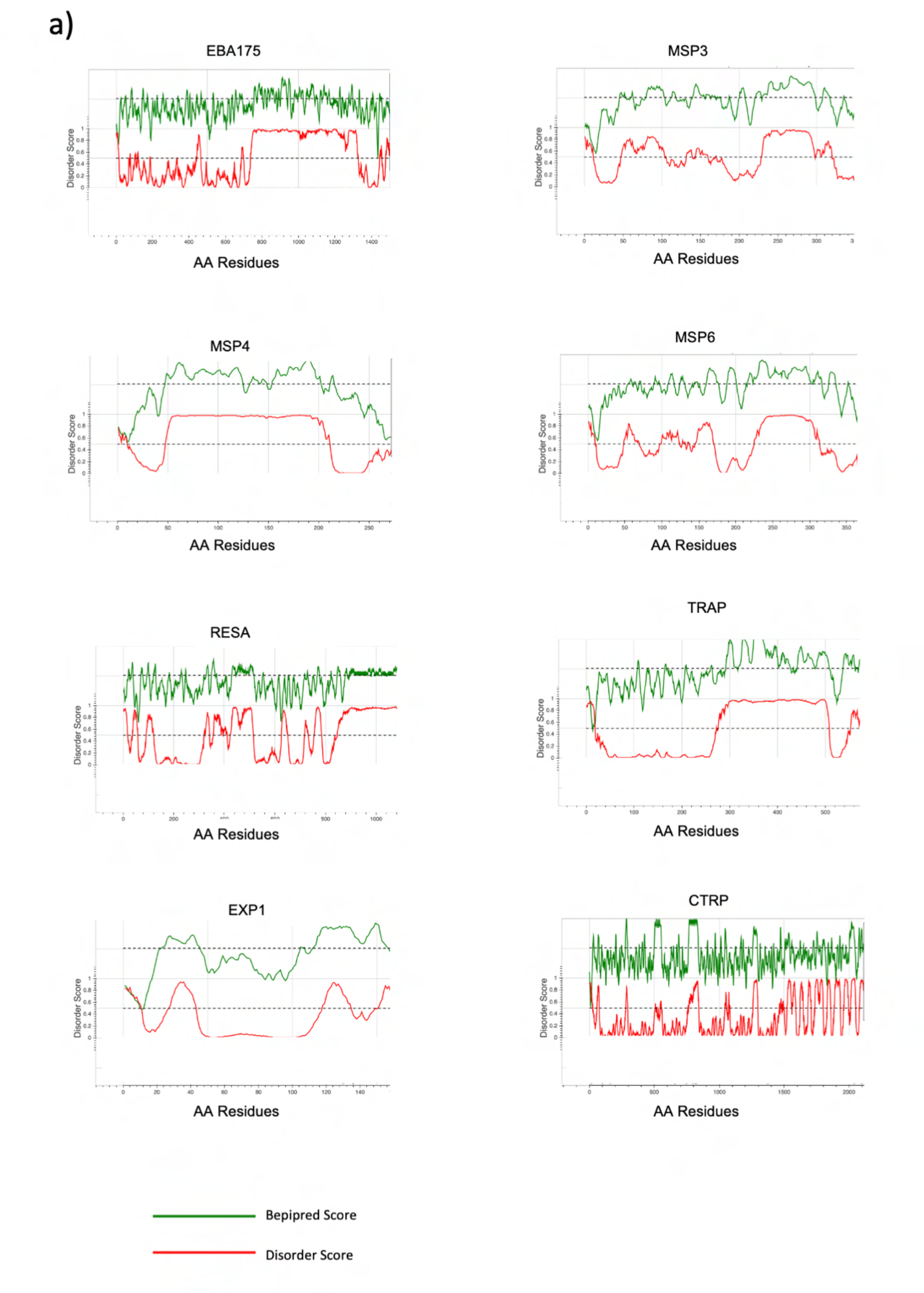
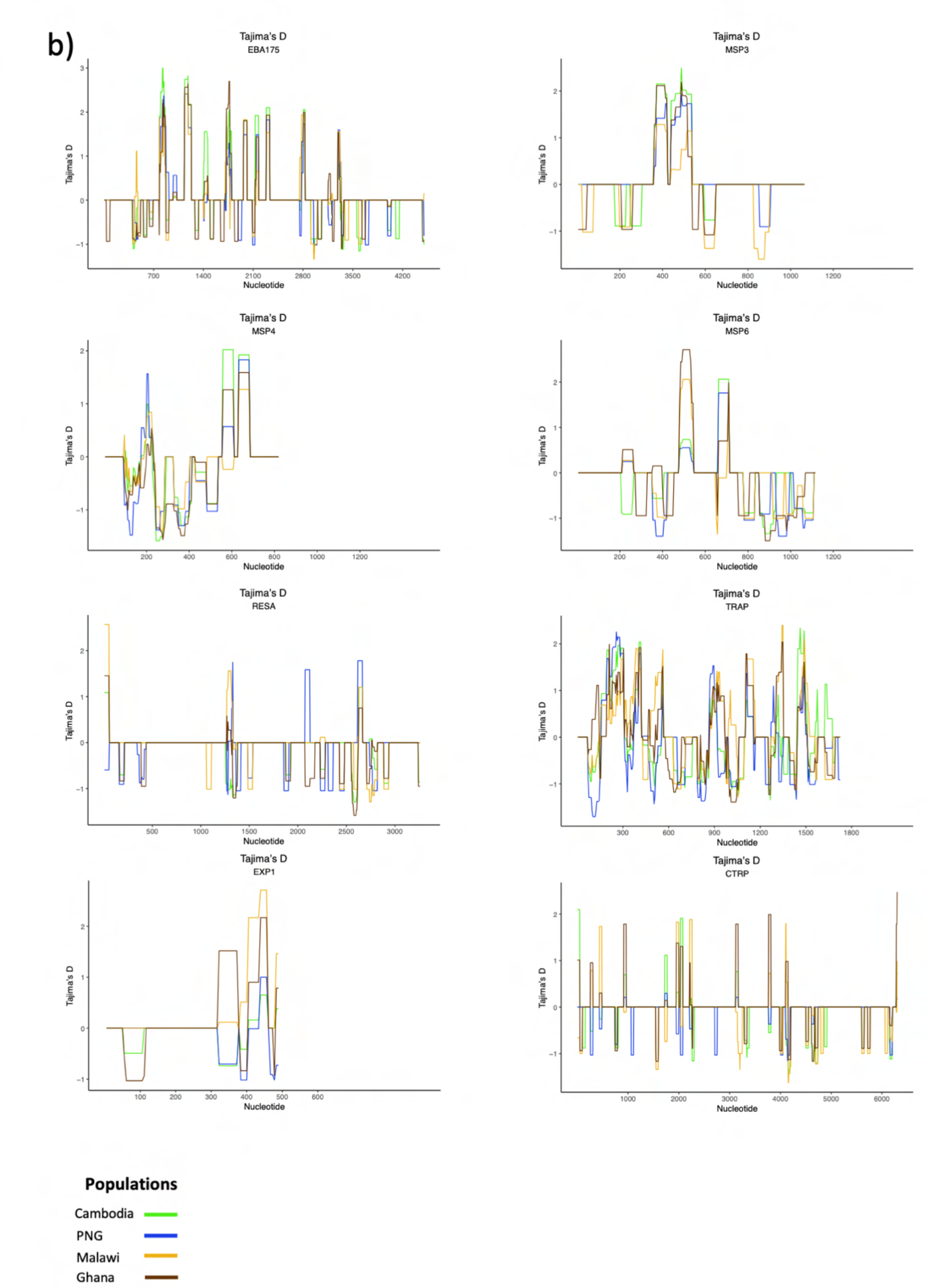
Selection of disordered proteins in Asia-Pacific and African regions. a) Computational predictions of protein disorder and B-cell epitopes in EBA175, MSP3, MSP4, MSP6, RESA, TRAP, EXP1 and CTRP. The green line represents the linear B-cell epitope mapping scores and the red line shows the protein disorder score, respectively. b) Tajima’s D statistics along the disordered antigens in samples from Cambodia, PNG, Malawi, and Ghana. It is calculated in the context of linear sequence level based on coding region with the sliding window approach (a window size of 50 bp and a step size of 5 bp). Nucleotide positions based on coding region are shown in the x-axis. Sample size for each respective population are as follows: Malawi (n= 106), Ghana (n=208), Cambodia (n=405), and PNG (n=108).

**Figure S11.**
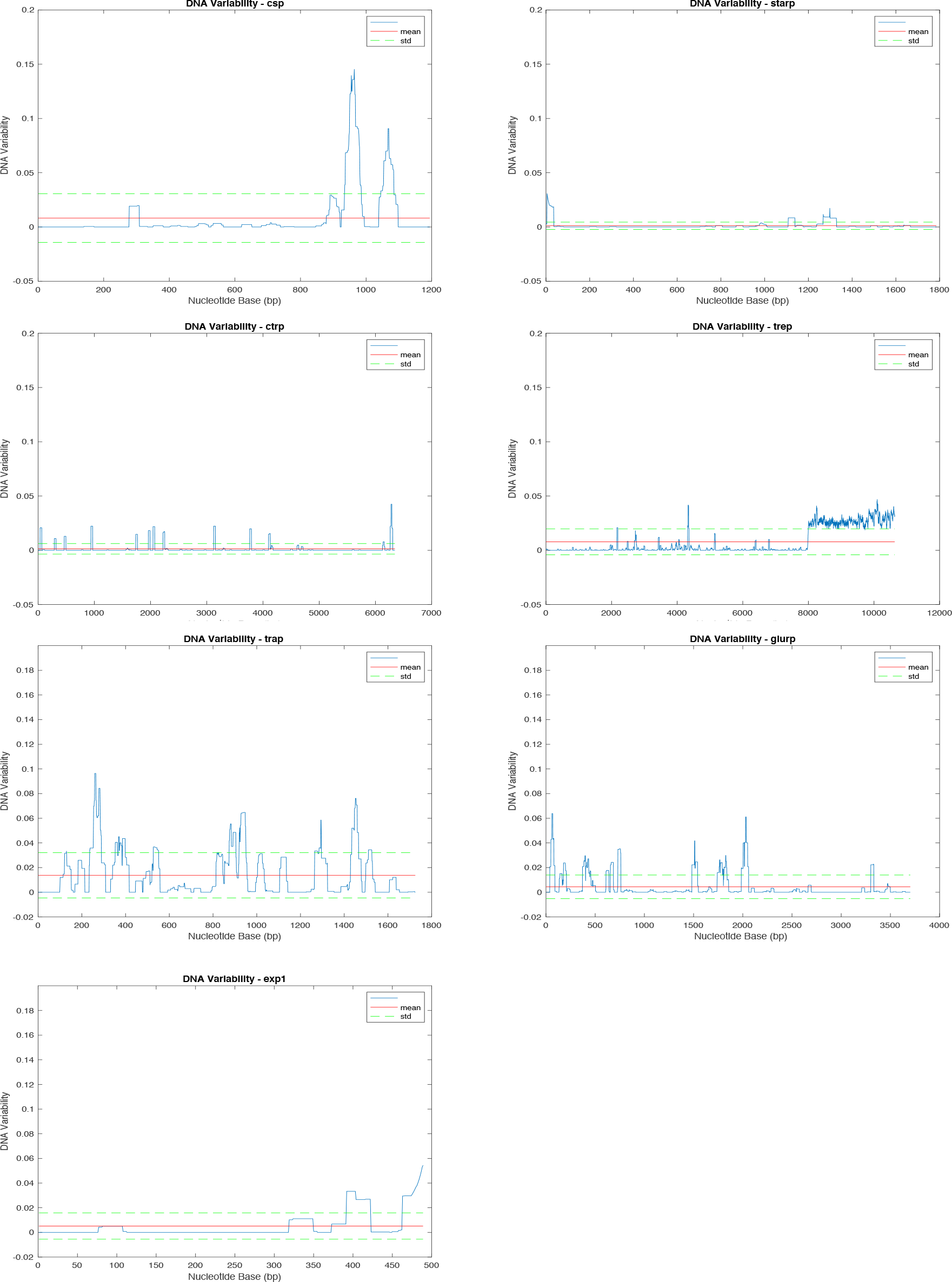
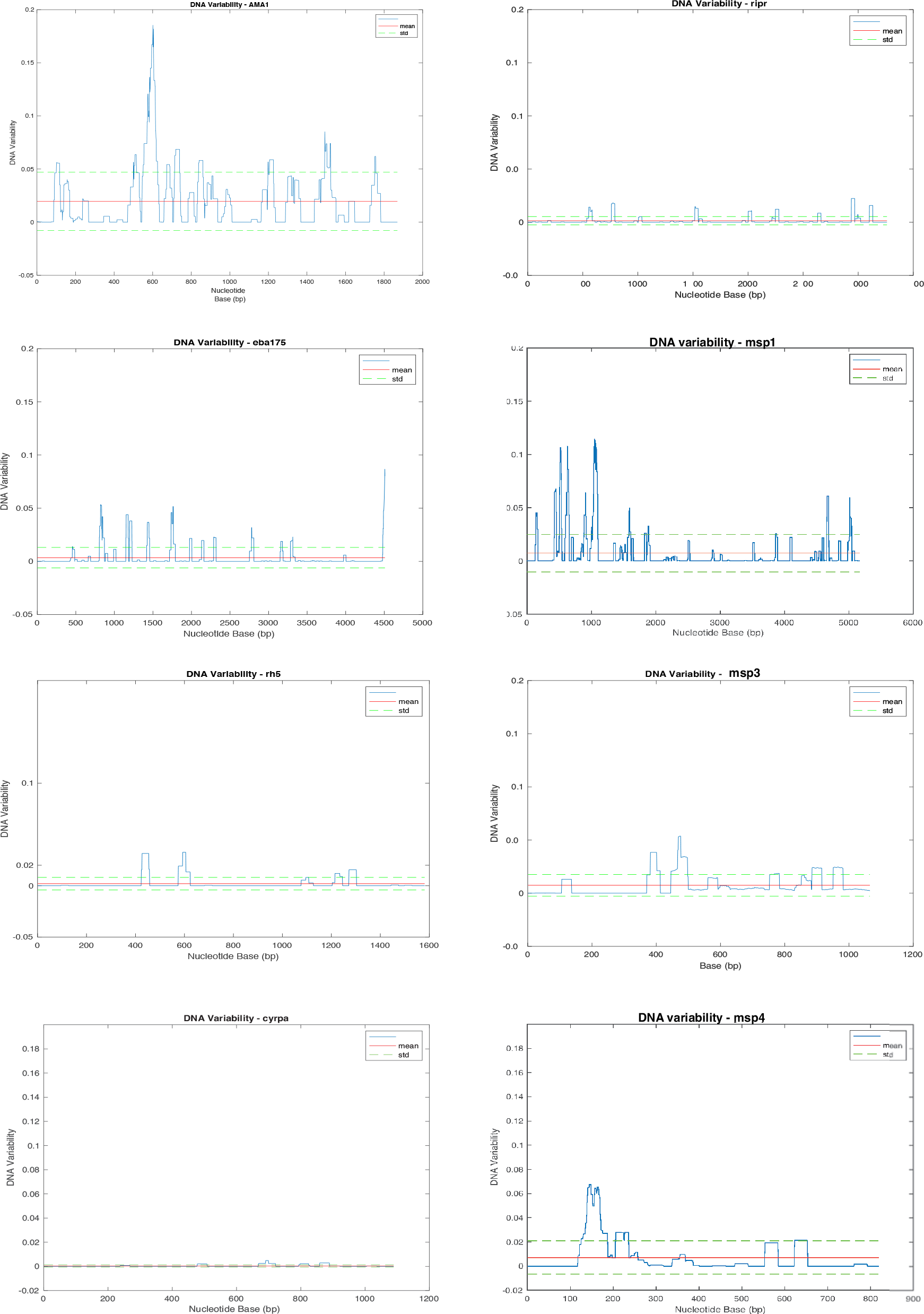
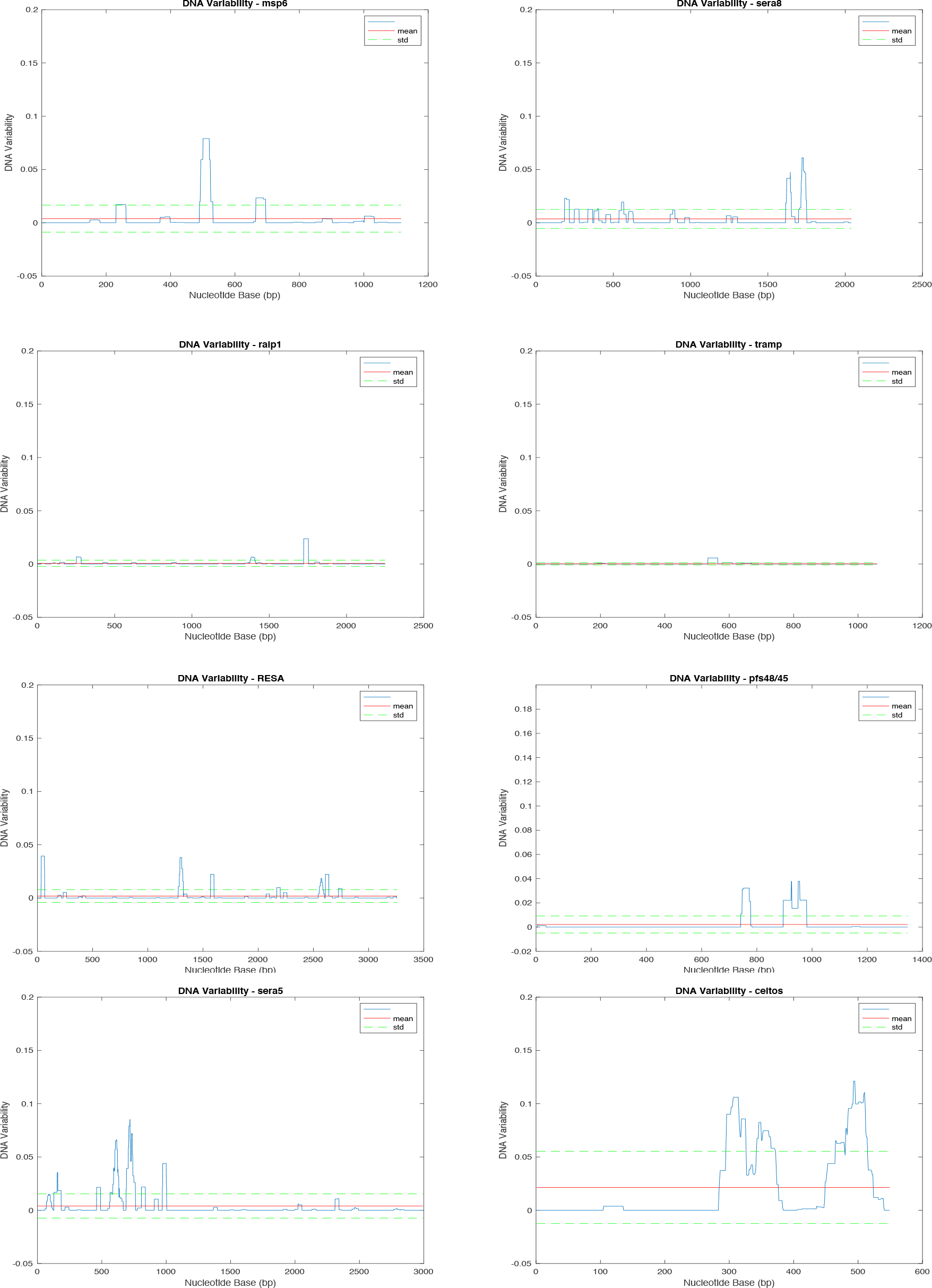
Nucleotide sequence variability of all antigens included in the study based on all available sequences. Sliding window analysis of sequence variability was calculated using algorithm from Proutski and Holmes et al (1997)^1^ implemented in MBEToolbox^2^ using default parameters on MATLAB (version R2020a). Mean (red line), and standard deviation (green dotted line) within each antigen are shown. Nucleotide positions based on coding region are shown in the x-axis.

